# Sex-specific risks for cardiovascular disease across the glycaemic spectrum: a population-based cohort study using the UK Biobank

**DOI:** 10.1101/2023.03.16.23287310

**Authors:** Christopher T Rentsch, Victoria Garfield, Rohini Mathur, Sophie V Eastwood, Liam Smeeth, Nish Chaturvedi, Krishnan Bhaskaran

## Abstract

**Background:** We sought to examine sex-specific risks for incident cardiovascular disease (CVD) across the full glycaemic spectrum.

**Methods:** Using data from UK Biobank, we categorised participants’ glycosylated haemoglobin (HbA1c) at baseline as low-normal (<35 mmol/mol), normal (35-41 mmol/mol), pre-diabetes (42-47 mmol/mol), undiagnosed diabetes (≥48 mmol/mol), or diagnosed diabetes. Our outcomes were coronary artery disease (CAD), atrial fibrillation, deep vein thrombosis (DVT), pulmonary embolism (PE), stroke, heart failure, and a composite outcome of any CVD. Cox regression estimated sex-specific associations between HbA1c and each outcome, sequentially adjusting for socio-demographic, lifestyle, and clinical characteristics.

**Findings:** Among 427,435 people, CVD rates were 16.9 and 9.1 events/1000 person-years for men and women, respectively. Both men and women with pre-diabetes, undiagnosed diabetes, and, more markedly, diagnosed diabetes were at higher risks of CVD than those with normal HbA1c, with relative increases more pronounced in women than men. Age-adjusted HRs for pre-diabetes and undiagnosed diabetes ranged from 1.30 to 1.47; HRs for diagnosed diabetes were 1.55 (1.49-1.61) in men and 2.00 (1.89-2.12) in women (p-interaction<0.0001). Excess risks attenuated and were more similar between men and women after adjusting for clinical and lifestyle factors particularly obesity and antihypertensive or statin use (fully adjusted HRs for diabetes: 1.06 [1.02-1.11] and 1.17 [1.10-1.24], respectively).

**Interpretation:** Excess risks in men and women were largely explained by modifiable factors, and could be ameliorated by attention to weight reduction strategies and greater use of antihypertensive and statin medications. Addressing these risk factors could reduce sex disparities in glycaemia-related risks of CVD.

**Funding:** Diabetes UK (#15/0005250) and British Heart Foundation (SP/16/6/32726)

**Research in context:** *Evidence before this study:* It has long been asserted that men and women with diabetes have differential risks of cardiovascular disease (CVD), but it is unclear which risk factors drive these sex differences, and whether men or women with moderately elevated glycosylated haemoglobin (HbA1c) below the threshold for diabetes are also at increased risk of CVD. We searched MEDLINE and PubMed on 15 March 2023 for studies evaluating sex differences in the risk of CVD across the glycaemic spectrum. The keywords “ (sex difference* OR sex disparit* OR sex-strat* OR sex-specific) AND (glycaemia OR glycemia OR glycosylated OR hemoglobin OR haemoglobin) AND (non-diabetes OR non-diabetic) AND (cardiovascular) AND (rate OR hazard OR odds OR risk)” were used and results were filtered to articles with an abstract available in English. 33 papers were identified and all 33 were eligible for screening; none reported sex-stratified associations of CVD risk across the full glycaemic spectrum. Previous studies have suggested a J-shaped curve in the relationship between HbA1c and outcomes including CVD and all-cause mortality, with individuals with low-normal HbA1c at excess risk compared to normal HbA1c. However, these studies were limited in sample size, combined people with controlled diabetes with those without diabetes, and did not analyse individual CVD outcomes. The present study leveraged UK Biobank data, which measured HbA1c on ∼500,000 men and women, regardless of diabetes status, presenting a unique opportunity to study sex disparities in the risk of CVD across the glycaemic spectrum.

*Added value of this study:* We uncovered novel insights around sex disparities in CVD risk across the glycaemic spectrum. Absolute CVD rates were higher in men than women at all levels of HbA1c. Both men and women with pre-diabetes, undiagnosed diabetes, and, more markedly, diagnosed diabetes were at higher risks of CVD than those with normal HbA1c, with relative increases in risk more pronounced in women than men. Both men and women with low-normal HbA1c had lower absolute rates of CVD than those with normal HbA1c. We extended previous evidence by showing most excess risk, and thereby differential relative risks between men and women, disappeared after accounting for lifestyle and clinical characteristics, namely measures of obesity and use of antihypertensive or lipid-lowering medications.

*Implications of all the available evidence:* This is the largest study to date to investigate sex differences in the risk of CVD across the glycaemic spectrum. While those with diagnosed diabetes carried the highest risk compared to those with normal HbA1c, men and women with pre-diabetes and undiagnosed diabetes were also at higher risk and those with low-normal HbA1c were at lower risk of CVD outcomes, highlighting the need for strategies to reduce risk of CVD across the glycaemic spectrum. Our findings suggest that excess risks in both men and women were largely explained by modifiable factors and could be ameliorated by attention to weight reduction strategies and greater use of antihypertensive and statin medications. Addressing these risk factors could reduce sex disparities in glycaemia-related risks of CVD.

## Background

With increasing global prevalence of hyperglycaemia and diabetes,^1–3^ understanding associations and mechanisms with its most important complication, cardiovascular disease (CVD), becomes increasingly important. It has long been asserted that men and women with diabetes have differential risks of CVD,^4–6^ but it is unclear which risk factors drive these sex differences, and whether men or women with moderately elevated glycaemia below the threshold for diabetes are also at increased risk of CVD. A study of the risk of adverse CVD outcomes across the full glycaemic spectrum among men and women with and without diagnosed diabetes could help further improve our understanding of underlying sex-specific mechanisms.

Previous meta-analyses have reported a 2.5- to 3.5-fold relative risk in the association between diabetes and CVD among women, compared to a 1.5- to 2.0- fold relative risk in men.^7–9^ The greater risk among women largely persists after multivariable adjustment, though interpretation is limited as studies are heterogeneous in design, outcome choice, and consideration of potential confounders. Recent evidence suggests sex differences may also vary by specific CVD outcome, with less of an excess observed in women for heart failure or stroke.^10^ The mechanisms through which these sex differences might operate have been suggested to include disparities in the identification, treatment, and control of cardiovascular risk factors.^6,11,12^ Understanding which of these factors are driving sex differences could help inform future evidence-based diabetes management guidelines.

We therefore aimed to examine sex-specific risks for six cardiovascular outcomes and the role of clinical and lifestyle characteristics across the full glycaemic spectrum.

## Methods

### Study design and population

We conducted an observational cohort study using data from UK Biobank,^13^ which includes 273,317 women and 229,081 men aged 40-69 recruited between 2006 and 2010 across England, Scotland, and Wales. Participants underwent baseline assessment capturing socio-demographic, lifestyle, and clinical factors, and gave blood samples for biomarker measurement. Participants also consented for linkage to hospital and death registry data. For this analysis, we excluded 1,836 participants with type 1 diabetes at baseline.

This study is reported according to the strengthening the reporting of observational studies in epidemiology (STROBE) and reporting of studies conducted using observational routinely collected health data (RECORD) guidelines (see **Supplementary Appendix**).

### Exposure, outcome, and follow-up

Glycosylated haemoglobin (HbA1c) was measured for all participants at recruitment, regardless of diabetes status. We excluded 35,999 (7%) participants who had missing HbA1c. We categorised participants at standard clinical cut-off points: low-normal (<35 mmol/mol or <5.5%), normal (35-41 mmol/mol or 5.5-5.9%), pre-diabetes (42-47 mmol/mol or 6.0-6.4%), undiagnosed diabetes (≥48 mmol/mol or ≥6.5%), or diagnosed diabetes defined by a diagnostic code or receipt of glucose-lowering medication.

We ascertained incidence of six CVD outcomes, namely coronary artery disease (CAD), atrial fibrillation, deep vein thrombosis (DVT), pulmonary embolism (PE), stroke, heart failure, and a composite outcome of any CVD. All outcomes were based on International Classification of Diseases – Tenth Edition (ICD-10) codes (**eTable 1**). We included primary and secondary diagnoses from the hospital registry and primary or contributing cause of death from the death registry. For the analysis of each outcome, we excluded individuals who had the respective event prior to baseline (e.g., those with CAD at baseline were excluded from the CAD analyses, those with atrial fibrillation at baseline were excluded from the atrial fibrillation analyses, etc).

Individuals were followed from the date of baseline assessment until earliest of: incident CVD, death, or end of data coverage (28 February 2021 in England & Scotland; 28 February 2018 in Wales).

### Covariates

Potential determinants of HbA1c and CVD were identified by reviewing existing literature and clinician consensus. We extracted the following variables from the baseline assessment data: socio-demographic factors (i.e., age, sex, ethnicity, index of multiple deprivation); lifestyle characteristics (i.e., smoking status, alcohol consumption, physical activity, body mass index, waist-hip ratio, and dietary intake); and clinical characteristics (i.e., total cholesterol, serum creatinine, C-reactive protein, diagnosed hypertension, use of antihypertensive medications or statins, and family history of CVD). We used a previously validated algorithm to identify individuals diagnosed with type 1 and 2 diabetes.^14^ Adverse waist-hip ratio was defined as ≥0.95 for men and ≥0.80 for women.^15^ Serum creatinine measurements were converted into estimated glomerular filtration rate (eGFR) using the chronic kidney disease epidemiology collaboration (CKD-EPI) equation.^16^ Smoking status, alcohol consumption, physical activity, dietary intake, diagnosed hypertension, use of antihypertensive medications or statins, and family history of cardiovascular disease were self-reported on surveys or during baseline interviews.

Missing data affected 37,277 (8%) of participants eligible for study inclusion. A large proportion of missingness was driven by lack of physical activity measures (n=10,754, 29%) and non-HbA1c laboratory measures (n=22,441, 60%). The majority (70%) of missing non-HbA1c laboratory measures at baseline resulted from laboratory reporting and data issues; therefore, we assumed these data were missing at random (MAR). Lifestyle measures were missing because participants responded, “prefers not to say,” thus these data were likely missing not at random (MNAR). We used complete case analysis because the overall level of missing data was low and a large proportion of missingness was likely to be MNAR. In this circumstance, although multiple imputation is not appropriate, a complete case analysis will be unbiased if, conditional on model covariates, missingness is independent of the outcome.^17^

### Statistical analysis

Incidence rates for each CVD outcome were estimated by HbA1c category and sex and age-standardised to the UK Biobank population. This was done by weighting the calculated incidence in the study population with the age and sex distribution in the full UK Biobank population, hence removing differences in incidence that could be attributed to those factors. Confidence intervals (CIs) were estimated using 500 bootstrap replications.

Hazard ratios (HR) and 95% CI for the association between HbA1c category and each CVD outcome were estimated using Cox proportional hazards models using days in study as the timescale. Each outcome-specific model was adjusted in five stages: 1) unadjusted; 2) age-adjusted; 3) socio-demographic-adjusted; 4) socio-demographic and lifestyle-adjusted; and 5) fully adjusted, which additionally adjusted for clinical characteristics. Age, body mass index, waist-hip ratio, total cholesterol and C-reactive protein were all modelled continuously using 4-knot restricted cubic splines. The proportional hazards assumption for each HbA1c category was assessed by inspecting whether scaled Schoenfeld residuals were independent of time (**eFigure 1**); there were no clear violations.

We then identified which factors were responsible for the greatest attenuation in the associations with Hba1c category and any CVD. We iteratively adjusted for each variable starting with an unadjusted model and building to a fully adjusted model separately for men and women. Within each broad category of variables (i.e., socio-demographics, lifestyle factors, clinical characteristics), we identified individual variables that attenuated the most excess risk.

### Sensitivity analyses

First, we added angina to the outcome definition of CAD and ran a separate model for ischemic stroke only. Second, we excluded individuals with HbA1c <20 mmol/mol, which suggests chronic illness and increased mortality risk.^18^ Third, we assessed the potential for reverse causality by excluding events that occurred within the first 180 days. Fourth, we excluded individuals with diagnosed diabetes at baseline to remove the potential influence of individuals with exposure to glucose-lowering medications in the model.

## Results

### Cohort description

Of 502,398 participants, we excluded 1,836 (0.4%) with known type 1 diabetes and 35,999 (7.2%) with missing HbA1c (**Figure 1**). Of the remaining 464,712, a further 37,277 (8.0%) were excluded for having any missing data. The final analysis included 427,435 participants, including 195,752 (45.8%) men and 231,683 (54.2%) women.

**Figure 1.**
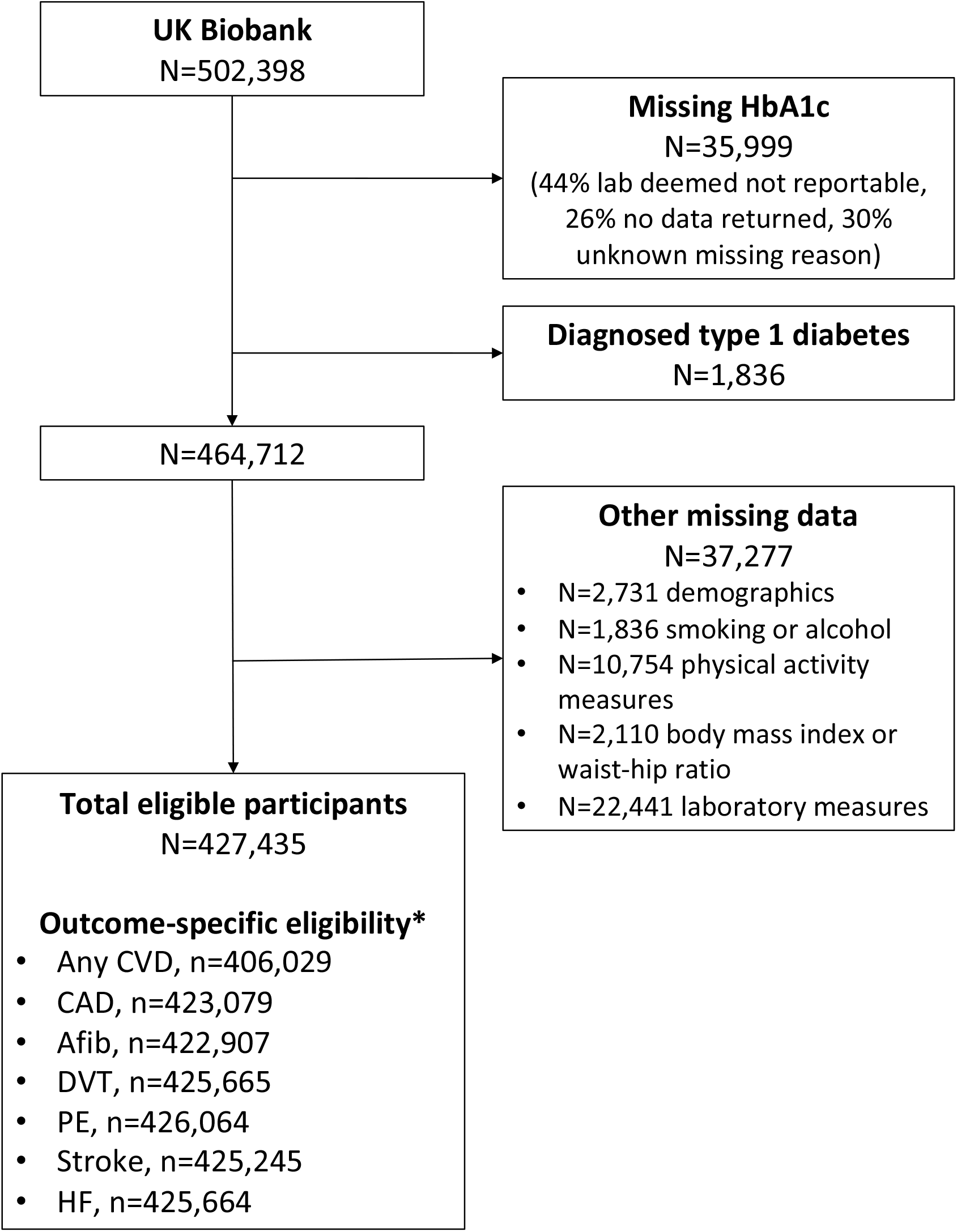
Study flow chart. *Excludes those with outcome on or before baseline. Notes: Demographics included age, sex, ethnicity, and index of multiple deprivation. Laboratory measures include total cholesterol, serum creatinine, and C-reactive protein.

Both men and women in higher HbA1c categories had higher body mass index, poorer renal function, greater prevalence of hypertension, and use of antihypertensive medications or statins compared to their counterparts with low-normal or normal HbA1c levels (**Table 1**). In keeping with the highest statin use, total cholesterol levels were lowest among those with diagnosed diabetes. Notably, the most favourable profiles were observed among participants in the low-normal HbA1c category.

**Table 1.** Cohort characteristics by sex and HbA1c category

|  | (A) Men |  |  |  |  |
| --- | --- | --- | --- | --- | --- |
|  | Low-normal<br>HbA1c | Normal HbA1c | Pre-diabetes | Undiagnosed<br>diabetes | Diagnosed<br>diabetes |
| Sample size, n | 90,745<br>(100.0) | 83,820<br>(100.0) | 6,873 (100.0) | 1,872 (100.0) | 12,442<br>(100.0) |
| **Socio-demographics** |  |  |  |  |  |
| Age, years (median (IQR)) | 56 (48-62) | 59 (52-64) | 61 (55-65) | 60 (53-64) | 62 (56-66) |
| Ethnicity |  |  |  |  |  |
| White European | 88,253 (97.3) | 79,499 (94.8) | 6,060 (88.2) | 1,635 (87.3) | 11,161 (89.7) |
| South Asian | 737 (0.8) | 1,512 (1.8) | 282 (4.1) | 91 (4.9) | 626 (5.0) |
| African Caribbean | 574 (0.6) | 1,124 (1.3) | 260 (3.8) | 68 (3.6) | 266 (2.1) |
| Mixed or other | 1,181 (1.3) | 1,685 (2.0) | 271 (3.9) | 78 (4.2) | 389 (3.1) |
| Index of multiple deprivation |  |  |  |  |  |
| 1 (least deprived) | 19,704 (21.7) | 17,209 (20.5) | 1,138 (16.6) | 287 (15.3) | 1,980 (15.9) |
| 2 | 19,120 (21.1) | 16,951 (20.2) | 1,253 (18.2) | 310 (16.6) | 2,137 (17.2) |
| 3 | 18,352 (20.2) | 16,841 (20.1) | 1,287 (18.7) | 336 (17.9) | 2,259 (18.2) |
| 4 | 17,697 (19.5) | 16,435 (19.6) | 1,398 (20.3) | 380 (20.3) | 2,524 (20.3) |
| 5 (most deprived) | 15,872 (17.5) | 16,384 (19.5) | 1,797 (26.1) | 559 (29.9) | 3,542 (28.5) |
| **Lifestyle characteristics** |  |  |  |  |  |
| Smoking status |  |  |  |  |  |
| Never | 60,439 (66.6) | 47,568 (56.8) | 3,122 (45.4) | 886 (47.3) | 5,888 (47.3) |
| Former | 22,101 (24.4) | 23,710 (28.3) | 2,432 (35.4) | 650 (34.7) | 5,082 (40.8) |
| Current | 8,205 (9.0) | 12,542 (15.0) | 1,319 (19.2) | 336 (17.9) | 1,472 (11.8) |
| Alcohol consumption |  |  |  |  |  |
| Daily or almost daily | 24,382 (26.9) | 21,552 (25.7) | 1,516 (22.1) | 396 (21.2) | 2,336 (18.8) |
| Three or four times a week | 25,779 (28.4) | 21,865 (26.1) | 1,447 (21.1) | 368 (19.7) | 2,415 (19.4) |
| Once or twice a week | 23,515 (25.9) | 21,794 (26.0) | 1,789 (26.0) | 490 (26.2) | 3,243 (26.1) |
| One to three times a month | 7,575 (8.3) | 7,382 (8.8) | 745 (10.8) | 199 (10.6) | 1,412 (11.3) |
| Special occasions only | 5,256 (5.8) | 6,156 (7.3) | 740 (10.8) | 202 (10.8) | 1,570 (12.6) |
| Never | 4,238 (4.7) | 5,071 (6.0) | 636 (9.3) | 217 (11.6) | 1,466 (11.8) |
| Days per week spent doing moderate or vigorous physical activity >10 mins |  |  |  |  |  |
| 0 | 9,471 (10.4) | 10,007 (11.9) | 1,152 (16.8) | 389 (20.8) | 2,436 (19.6) |
| 1-2 | 20,491 (22.6) | 17,818 (21.3) | 1,375 (20.0) | 414 (22.1) | 2,564 (20.6) |
| 3-7 | 60,783 (67.0) | 55,995 (66.8) | 4,346 (63.2) | 1,069 (57.1) | 7,442 (59.8) |
| Body mass index |  |  |  |  |  |
| Underweight | 227 (0.3) | 193 (0.2) | 10 (0.1) | 1 (0.1) | 11 (0.1) |
| Normal | 26,812 (29.5) | 19,569 (23.3) | 753 (11.0) | 118 (6.3) | 1,056 (8.5) |
| Overweight | 46,970 (51.8) | 41,911 (50.0) | 2,957 (43.0) | 653 (34.9) | 4,725 (38.0) |
| Obese | 16,736 (18.4) | 22,147 (26.4) | 3,153 (45.9) | 1,100 (58.8) | 6,650 (53.4) |
| Waist-hip ratio |  |  |  |  |  |
| Normal | 62,925 (69.3) | 49,011 (58.5) | 2,552 (37.1) | 519 (27.7) | 3,767 (30.3) |
| Adverse | 27,820 (30.7) | 34,809 (41.5) | 4,321 (62.9) | 1,353 (72.3) | 8,675 (69.7) |
| Number of times per week consuming processed meats |  |  |  |  |  |
| None | 5,324 (5.9) | 4,009 (4.8) | 337 (4.9) | 87 (4.6) | 637 (5.1) |
| <1 | 20,338 (22.4) | 17,576 (21.0) | 1,359 (19.8) | 332 (17.7) | 2,634 (21.2) |
| 1 | 27,292 (30.1) | 24,893 (29.7) | 1,955 (28.4) | 535 (28.6) | 3,585 (28.8) |
| 2-4 | 32,311 (35.6) | 31,707 (37.8) | 2,718 (39.5) | 751 (40.1) | 4,739 (38.1) |
| 5+ | 5,480 (6.0) | 5,635 (6.7) | 504 (7.3) | 167 (8.9) | 847 (6.8) |
| Number of times per day consuming fruit or vegetables |  |  |  |  |  |
| None | 1,354 (1.5) | 1,584 (1.9) | 181 (2.6) | 57 (3.0) | 205 (1.6) |
| 1-2 | 5,705 (6.3) | 6,008 (7.2) | 589 (8.6) | 178 (9.5) | 698 (5.6) |
| 3-4 | 16,244 (17.9) | 15,117 (18.0) | 1,206 (17.5) | 354 (18.9) | 1,797 (14.4) |
| 5+ | 67,442 (74.3) | 61,111 (72.9) | 4,897 (71.2) | 1,283 (68.5) | 9,742 (78.3) |
| **Clinical characteristics** |  |  |  |  |  |
| HbA1c, mmol/mol (median (IQR)) | 32.5 (30.8-33.8) | 37.1 (36.0-38.7) | 43.5 (42.6-44.9) | 53.2 (49.7-62.2) | 49.7 (43.1-58.3) |
| Total cholesterol, mmol/L (median (IQR)) | 5.5 (4.9-6.2) | 5.6 (4.8-6.3) | 5.3 (4.5-6.1) | 5.5 (4.6-6.3) | 4.2 (3.6-4.8) |
| eGFR, ml/min/1.73m<sup>2</sup> |  |  |  |  |  |
| ≥90 | 55,964 (61.7) | 45,633 (54.4) | 3,416 (49.7) | 1,172 (62.6) | 6,880 (55.3) |
| 60-89 | 33,495 (36.9) | 36,144 (43.1) | 3,147 (45.8) | 652 (34.8) | 4,757 (38.2) |
| <60 | 1,286 (1.4) | 2,043 (2.4) | 310 (4.5) | 48 (2.6) | 805 (6.5) |
| C-reactive protein, mg/L (median (IQR)) | 1.1 (0.6-2.1) | 1.4 (0.7-2.8) | 2.1 (1.1-4.2) | 2.8 (1.5-5.2) | 1.6 (0.8-3.1) |
| Hypertension | 21,801 (24.0) | 26,214 (31.3) | 3,081 (44.8) | 800 (42.7) | 8,349 (67.1) |
| On antihypertensive medication | 15,467 (17.0) | 21,698 (25.9) | 2,861 (41.6) | 641 (34.2) | 8,595 (69.1) |
| On statin | 10,708 (11.8) | 18,784 (22.4) | 2,601 (37.8) | 543 (29.0) | 9,346 (75.1) |
| Family history of cardiovascular disease | 47,331 (52.2) | 46,263 (55.2) | 3,948 (57.4) | 1,012 (54.1) | 7,569 (60.8) |

|  | (B) Women |  |  |  |  |
| --- | --- | --- | --- | --- | --- |
|  | Low-normal<br>HbA1c | Normal HbA1c | Pre-diabetes | Undiagnosed<br>diabetes | Diagnosed<br>diabetes |
| Sample size, n | 110,304<br>(100.0) | 105,533<br>(100.0) | 7,514<br>(100.0) | 1,216<br>(100.0) | 7,116<br>(100.0) |
| **Socio-demographics** |  |  |  |  |  |
| Age, years (median (IQR)) | 54 (47-61) | 60 (54-64) | 62 (57-65) | 60 (55-64) | 61 (56-65) |
| Ethnicity |  |  |  |  |  |
| White European | 106,805 (96.8) | 100,334 (95.1) | 6,576 (87.5) | 997 (82.0) | 6,192 (87.0) |
| South Asian | 748 (0.7) | 1,374 (1.3) | 236 (3.1) | 82 (6.7) | 343 (4.8) |
| African Caribbean | 873 (0.8) | 1,468 (1.4) | 390 (5.2) | 78 (6.4) | 282 (4.0) |
| Mixed or other | 1,878 (1.7) | 2,357 (2.2) | 312 (4.2) | 59 (4.9) | 299 (4.2) |
| Index of multiple deprivation |  |  |  |  |  |
| 1 (least deprived) | 23,176 (21.0) | 21,546 (20.4) | 1,316 (17.5) | 181 (14.9) | 978 (13.7) |
| 2 | 22,816 (20.7) | 21,574 (20.4) | 1,332 (17.7) | 186 (15.3) | 1,151 (16.2) |
| 3 | 22,675 (20.6) | 21,737 (20.6) | 1,398 (18.6) | 222 (18.3) | 1,340 (18.8) |
| 4 | 22,419 (20.3) | 21,028 (19.9) | 1,620 (21.6) | 265 (21.8) | 1,536 (21.6) |
| 5 (most deprived) | 19,218 (17.4) | 19,648 (18.6) | 1,848 (24.6) | 362 (29.8) | 2,111 (29.7) |
| **Lifestyle characteristics** |  |  |  |  |  |
| Smoking status |  |  |  |  |  |
| Never | 81,504 (73.9) | 74,067 (70.2) | 4,949 (65.9) | 824 (67.8) | 4,726 (66.4) |
| Former | 20,846 (18.9) | 20,868 (19.8) | 1,523 (20.3) | 252 (20.7) | 1,774 (24.9) |
| Current | 7,954 (7.2) | 10,598 (10.0) | 1,042 (13.9) | 140 (11.5) | 616 (8.7) |
| Alcohol consumption |  |  |  |  |  |
| Daily or almost daily | 20,314 (18.4) | 16,278 (15.4) | 801 (10.7) | 72 (5.9) | 536 (7.5) |
| Three or four times a week | 25,972 (23.5) | 20,639 (19.6) | 960 (12.8) | 135 (11.1) | 643 (9.0) |
| Once or twice a week | 29,768 (27.0) | 26,868 (25.5) | 1,744 (23.2) | 247 (20.3) | 1,326 (18.6) |
| One to three times a month | 13,664 (12.4) | 14,414 (13.7) | 1,112 (14.8) | 201 (16.5) | 996 (14.0) |
| Special occasions only | 13,173 (11.9) | 16,805 (15.9) | 1,744 (23.2) | 311 (25.6) | 2,060 (28.9) |
| Never | 7,413 (6.7) | 10,529 (10.0) | 1,153 (15.3) | 250 (20.6) | 1,555 (21.9) |
| Days per week spent doing moderate or vigorous physical activity >10 mins |  |  |  |  |  |
| 0 | 13,679 (12.4) | 13,426 (12.7) | 1,247 (16.6) | 248 (20.4) | 1,431 (20.1) |
| 1-2 | 24,748 (22.4) | 22,069 (20.9) | 1,494 (19.9) | 246 (20.2) | 1,441 (20.3) |
| 3-7 | 71,877 (65.2) | 70,038 (66.4) | 4,773 (63.5) | 722 (59.4) | 4,244 (59.6) |
| Body mass index |  |  |  |  |  |
| Underweight | 902 (0.8) | 791 (0.7) | 32 (0.4) | 1 (0.1) | 14 (0.2) |
| Normal | 51,026 (46.3) | 37,839 (35.9) | 1,315 (17.5) | 98 (8.1) | 715 (10.0) |
| Overweight | 40,211 (36.5) | 40,185 (38.1) | 2,568 (34.2) | 347 (28.5) | 1,936 (27.2) |
| Obese | 18,165 (16.5) | 26,718 (25.3) | 3,599 (47.9) | 770 (63.3) | 4,451 (62.5) |
| Waist-hip ratio |  |  |  |  |  |
| Normal | 56,636 (51.3) | 40,015 (37.9) | 1,289 (17.2) | 95 (7.8) | 706 (9.9) |
| Adverse | 53,668 (48.7) | 65,518 (62.1) | 6,225 (82.8) | 1,121 (92.2) | 6,410 (90.1) |
| Number of times per week consuming processed meats |  |  |  |  |  |
| None | 14,624 (13.3) | 12,747 (12.1) | 780 (10.4) | 126 (10.4) | 720 (10.1) |
| <1 | 42,936 (38.9) | 40,367 (38.3) | 2,649 (35.3) | 422 (34.7) | 2,461 (34.6) |
| 1 | 31,084 (28.2) | 30,424 (28.8) | 2,294 (30.5) | 349 (28.7) | 2,119 (29.8) |
| 2-4 | 19,754 (17.9) | 20,166 (19.1) | 1,637 (21.8) | 287 (23.6) | 1,628 (22.9) |
| 5+ | 1,906 (1.7) | 1,829 (1.7) | 154 (2.0) | 32 (2.6) | 188 (2.6) |
| Number of times per day consuming fruit or vegetables |  |  |  |  |  |
| None | 746 (0.7) | 819 (0.8) | 96 (1.3) | 16 (1.3) | 64 (0.9) |
| 1-2 | 3,379 (3.1) | 3,318 (3.1) | 289 (3.8) | 55 (4.5) | 230 (3.2) |
| 3-4 | 12,718 (11.5) | 11,612 (11.0) | 932 (12.4) | 159 (13.1) | 695 (9.8) |
| 5+ | 93,461 (84.7) | 89,784 (85.1) | 6,197 (82.5) | 986 (81.1) | 6,127 (86.1) |
| **Clinical characteristics** |  |  |  |  |  |
| HbA1c, mmol/mol (median (IQR)) | 32.6 (30.9-33.8) | 37.1 (36.0-38.6) | 43.3 (42.5-44.6) | 51.6 (49.4-57.9) | 49.6 (43.3-58.0) |
| Total cholesterol, mmol/L (median (IQR)) | 5.7 (5.0-6.4) | 6.0 (5.3-6.8) | 5.8 (5.1-6.7) | 5.9 (5.1-6.8) | 4.6 (4.0-5.3) |
| eGFR, ml/min/1.73m<sup>2</sup> |  |  |  |  |  |
| ≥90 | 71,799 (65.1) | 57,648 (54.6) | 3,676 (48.9) | 772 (63.5) | 4,032 (56.7) |
| 60-89 | 37,003 (33.5) | 45,125 (42.8) | 3,490 (46.4) | 403 (33.1) | 2,616 (36.8) |
| <60 | 1,502 (1.4) | 2,760 (2.6) | 348 (4.6) | 41 (3.4) | 468 (6.6) |
| C-reactive protein, mg/L (median (IQR)) | 1.1 (0.5-2.3) | 1.5 (0.7-3.2) | 2.8 (1.3-5.7) | 4.3 (2.3-8.0) | 2.5 (1.1-5.1) |
| Hypertension | 19,612 (17.8) | 27,515 (26.1) | 3,159 (42.0) | 511 (42.0) | 4,629 (65.1) |
| On antihypertensive medication | 13,040 (11.8) | 20,721 (19.6) | 2,708 (36.0) | 424 (34.9) | 4,540 (63.8) |
| On statin | 5,533 (5.0) | 13,456 (12.8) | 2,021 (26.9) | 304 (25.0) | 5,064 (71.2) |
| Family history of cardiovascular disease | 60,735 (55.1) | 64,721 (61.3) | 4,915 (65.4) | 748 (61.5) | 4,758 (66.9) |

*Abbreviations:* IQR, interquartile range; HbA1c, glycosylated haemoglobin; eGFR, estimated glomerular filtration rate
*Notes:* Categories were defined by baseline HbA1c levels as follows: low-normal (<35 mmol/mol or <5.5%), normal (35-41 mmol/mol or 5.5-5.9%), pre-diabetes (42-47 mmol/mol or 6.0-6.4%), undiagnosed diabetes ( ≥ 48 mmol/mol or ≥ 6.5%), or diagnosed diabetes defined by a diagnostic code or receipt of glucose-lowering medication.

Women with diagnosed diabetes had a more marked excess of adverse risk factors than men (**Table 1**). This was particularly true for adverse waist-hip ratio (90.1% for women versus 69.7% for men), obesity (62.5% for women versus 53.4% for men), lower use of antihypertensive medications (63.8% for women versus 69.1% for men), and statins (71.2% for women versus 75.1% for men). Similarly, use of these medications was lower in women with normal HbA1c compared to men (antihypertensives: 19.6% for women versus 25.9% for men; statins: 12.8% for women versus 22.4% for men).

### Age-standardised incidence rates

Over a median 11.8 years of follow-up, we observed 51,288 incident cardiovascular events. Age-standardised incidence rates of any CVD were 16.9 and 9.1 events per 1000 person-years (PY) for men and women, respectively. Among men, CVD rates were similar in those with pre-diabetes (21.8/1000 PY) and undiagnosed (21.7/1000 PY) and diagnosed diabetes (26.8/1000 PY), and markedly elevated compared to those with normal HbA1c (16.5/1000 PY; **Table 2**). In contrast, CVD rates were lower in those with low-normal HbA1c (14.0/1000 PY) compared to normal HbA1c. Patterns were similar among women.

**Table 2.**
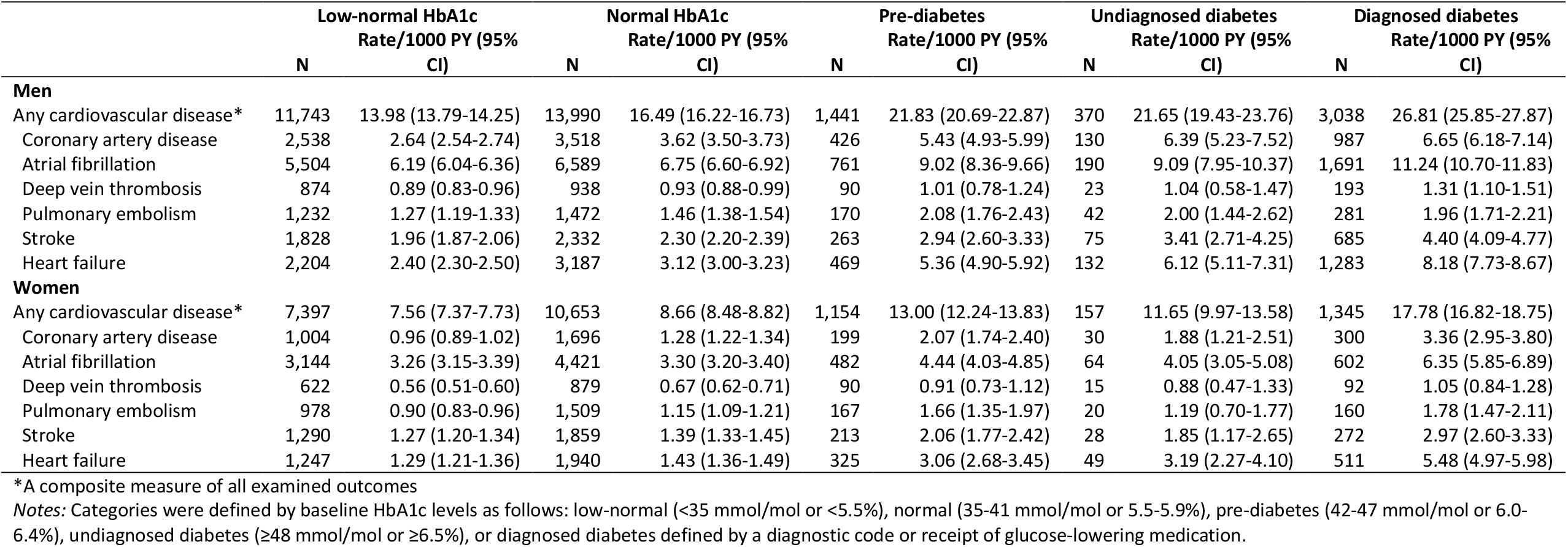
Age-standardised incidence rates by sex and HbA1c category

### HbA1c category and CVD risk

Age-adjusted relative associations between diabetes and any CVD were stronger for women than men (HR 2.00, 95%CI 1.89-2.12 for women; HR 1.55, 95%CI 1.49-1.61 for men; p-interaction<0.0001; **Figure 2** and **eTable 2**). Compared to those with normal HbA1c, both women and men with pre-diabetes or undiagnosed diabetes were also at elevated risk of CVD (HR 1.47, 95% CI 1.38-1.56 for pre-diabetes and HR 1.33, 95% CI 1.14-1.56 for undiagnosed diabetes among women; HR 1.30, 95% CI 1.24-1.38 for pre-diabetes and HR 1.31, 95% CI 1.18-1.45 for undiagnosed diabetes among men). In addition, both women and men with low-normal HbA1c were at decreased risk of CVD (HR 0.86, 95% CI 0.84-0.98 for women; HR 0.86, 95% CI 0.84-0.88 for men). Associations attenuated with additional adjustment for socio-demographic, lifestyle, and clinical characteristics. However, both women and men with diagnosed diabetes remained at elevated risk for CVD (HR 1.17, 95% CI 1.10-1.24 for women; HR 1.06, 95% CI 1.02-1.11 for men; p-interaction=0.0387).

**Figure 2.**
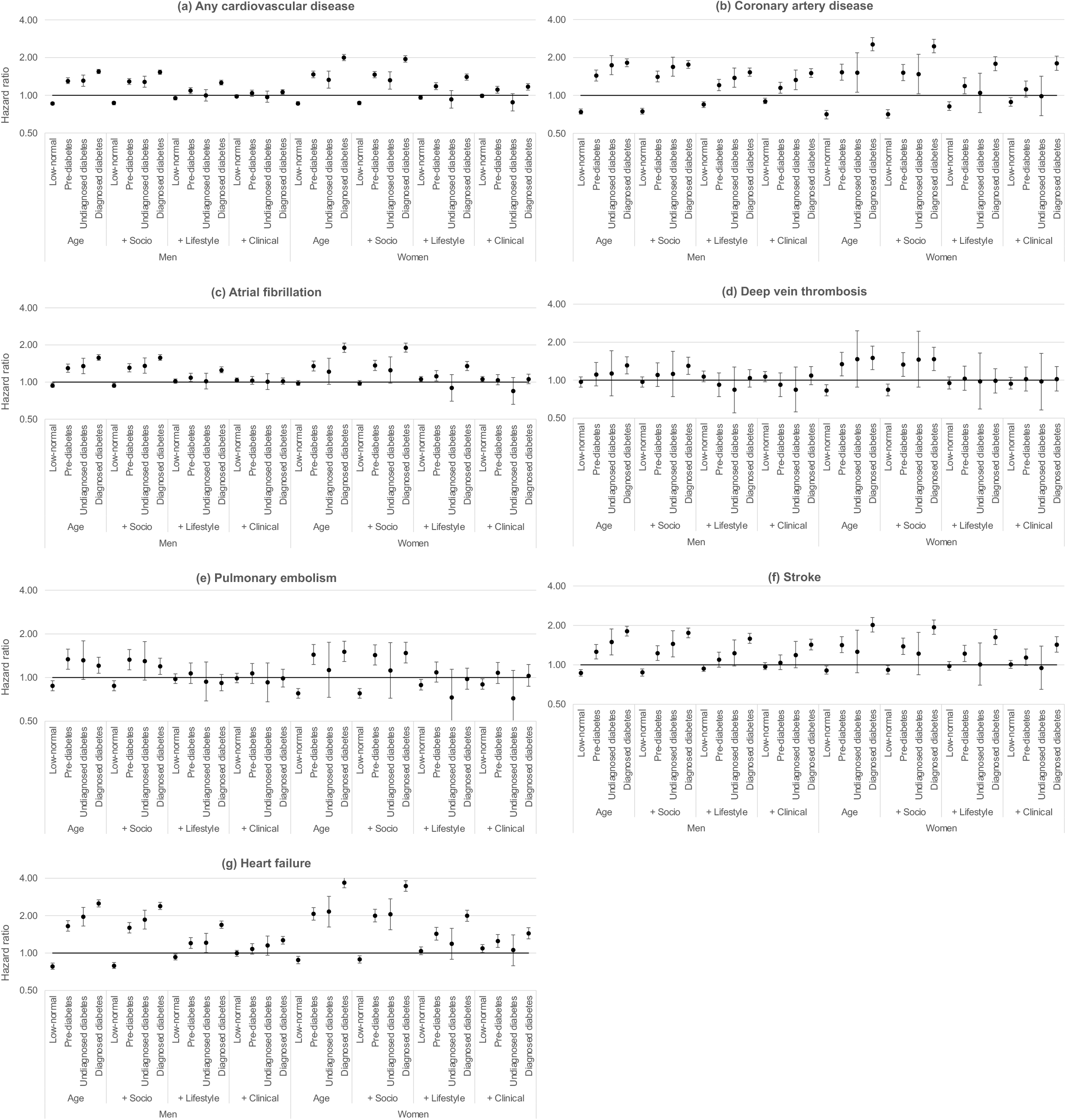
Sex-specific, fully adjusted associations between glycated haemoglobin (HbA1c) and six cardiovascular diseases. *A composite measure of all examined outcomes. Notes: Sex-specific hazard ratios from Cox proportional hazards models, sequentially adjusted for age at study entry, socio-demographics (i.e., ethnicity and deprivation), lifestyle factors (i.e., smoking status, alcohol consumption, physical activity, body mass index, waist-hip ratio, processed meat and fruit and vegetable intake), and clinical characteristics (i.e., total cholesterol, estimated glomerular filtration rate, C-reactive protein, diagnosed hypertension, use of antihypertensive medication or statins, and family history of cardiovascular disease). Categories defined as follows: low-normal (<35 mmol/mol or <5.5%), pre-diabetes (42-47 mmol/mol or 6.0-6.4%), undiagnosed diabetes (≥48 mmol/mol or ≥6.5%), or diagnosed diabetes defined by a diagnostic code or receipt of glucose-lowering medication. Reference group: normal HbA1c (35-41 mmol/mol or 5.5-5.9%).

In fully adjusted models, men and women with diagnosed diabetes were at greater risk for CAD (HR 1.51, 95% CI 1.39-1.63 for men and HR 1.80, 95% CI 1.58-2.05 for women; p-interaction=0.0540), stroke (HR 1.43, 95% CI 1.30-1.57 for men and HR 1.43, 95% CI 1.25-1.64 for women; p-interaction=0.6596), and heart failure (HR 1.27, 95% CI 1.18-1.36 for men and HR 1.44, 95% CI 1.30-1.60 for women; p-interaction=0.0678) compared to their counterparts with normal HbA1c (**Figure 2** and **eTable 2**). Men and women with low-normal HbA1c were at decreased risk of CAD (HR 0.90, 95% CI 0.86-0.95 for men and HR 0.89, 95% CI 0.82-0.96 for women). In addition, men with pre-diabetes (HR 1.15, 95%CI 1.04-1.27) or undiagnosed diabetes (HR 1.33, 95%CI 1.11-1.59) were also at elevated risk of CAD; these associations were not observed among women. Similar patterns of associations between HbA1c category and risk of atrial fibrillation, DVT, and PE were observed in age-adjusted models, with relative risks higher among women than men; however, no strong evidence of associations or sex differences were observed among men or women for these outcomes after full adjustment.

### Identifying factors most responsible for attenuating excess risk

After already accounting for age, further adjustment for socio-demographic variables did not materially alter the greater risk of any CVD associated with pre-diabetes, undiagnosed diabetes, or diagnosed diabetes among men or women (**Table 3**). However, adjustment for both lifestyle factors, and, separately, clinical factors, markedly reduced excess risk. Inspection of the role of individual factors revealed the following: accounting for body mass index, waist-hip ratio, and use of antihypertensive medications or statins had the greatest impact on attenuating the increased risk of any CVD associated with elevated HbA1c or diagnosed diabetes and decreased risk of any CVD associated with low-normal HbA1c. Excess risk mostly disappeared after adjustment for all factors, though there remained some evidence of differential risk between men and women in fully adjusted models.

**Table 3.**
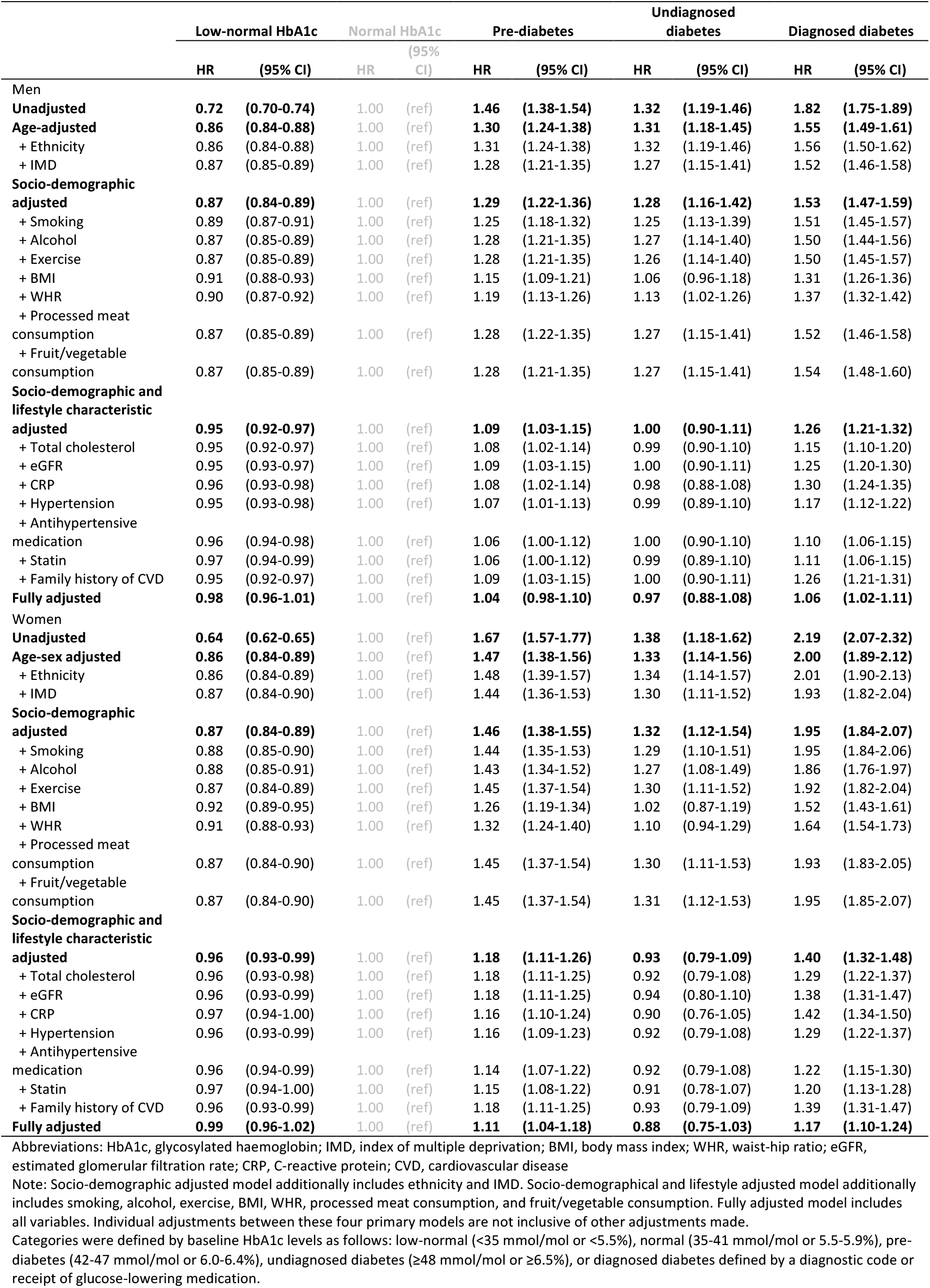
Individual adjustments for the associations between HbA1c category and any cardiovascular disease

|  | Low-normal HbA1c |  | Normal HbA1c |  | Pre-diabetes |  | Undiagnosed diabetes |  | Diagnosed diabetes |  |
| --- | --- | --- | --- | --- | --- | --- | --- | --- | --- | --- |
|  | HR | (95% CI) | HR | (95% CI) | HR | (95% CI) | HR | (95% CI) | HR | (95% CI) |
| **Men** |  |  |  |  |  |  |  |  |  |  |
| **Unadjusted** | **0.72** | **(0.70-0.74)** | 1.00 | (ref) | **1.46** | **(1.38-1.54)** | **1.32** | **(1.19-1.46)** | **1.82** | **(1.75-1.89)** |
| **Age-adjusted** | **0.86** | **(0.84-0.88)** | 1.00 | (ref) | **1.30** | **(1.24-1.38)** | **1.31** | **(1.18-1.45)** | **1.55** | **(1.49-1.61)** |
| + Ethnicity | 0.86 | (0.84-0.88) | 1.00 | (ref) | 1.31 | (1.24-1.38) | 1.32 | (1.19-1.46) | 1.56 | (1.50-1.62) |
| + IMD | 0.87 | (0.85-0.89) | 1.00 | (ref) | 1.28 | (1.21-1.35) | 1.27 | (1.15-1.41) | 1.52 | (1.46-1.58) |
| **Socio-demographic adjusted** | **0.87** | **(0.84-0.89)** | 1.00 | (ref) | **1.29** | **(1.22-1.36)** | **1.28** | **(1.16-1.42)** | **1.53** | **(1.47-1.59)** |
| + Smoking | 0.89 | (0.87-0.91) | 1.00 | (ref) | 1.25 | (1.18-1.32) | 1.25 | (1.13-1.39) | 1.51 | (1.45-1.57) |
| + Alcohol | 0.87 | (0.85-0.89) | 1.00 | (ref) | 1.28 | (1.21-1.35) | 1.27 | (1.14-1.40) | 1.50 | (1.44-1.56) |
| + Exercise | 0.87 | (0.85-0.89) | 1.00 | (ref) | 1.28 | (1.21-1.35) | 1.26 | (1.14-1.40) | 1.50 | (1.45-1.57) |
| + BMI | 0.91 | (0.88-0.93) | 1.00 | (ref) | 1.15 | (1.09-1.21) | 1.06 | (0.96-1.18) | 1.31 | (1.26-1.36) |
| + WHR | 0.90 | (0.87-0.92) | 1.00 | (ref) | 1.19 | (1.13-1.26) | 1.13 | (1.02-1.26) | 1.37 | (1.32-1.42) |
| + Processed meat consumption | 0.87 | (0.85-0.89) | 1.00 | (ref) | 1.28 | (1.22-1.35) | 1.27 | (1.15-1.41) | 1.52 | (1.46-1.58) |
| + Fruit/vegetable consumption | 0.87 | (0.85-0.89) | 1.00 | (ref) | 1.28 | (1.21-1.35) | 1.27 | (1.15-1.41) | 1.54 | (1.48-1.60) |
| **Socio-demographic and lifestyle characteristic adjusted** | **0.95** | **(0.92-0.97)** | 1.00 | (ref) | **1.09** | **(1.03-1.15)** | **1.00** | **(0.90-1.11)** | **1.26** | **(1.21-1.32)** |
| + Total cholesterol | 0.95 | (0.92-0.97) | 1.00 | (ref) | 1.08 | (1.02-1.14) | 0.99 | (0.90-1.10) | 1.15 | (1.10-1.20) |
| + eGFR | 0.95 | (0.93-0.97) | 1.00 | (ref) | 1.09 | (1.03-1.15) | 1.00 | (0.90-1.11) | 1.25 | (1.20-1.30) |
| + CRP | 0.96 | (0.93-0.98) | 1.00 | (ref) | 1.08 | (1.02-1.14) | 0.98 | (0.88-1.08) | 1.30 | (1.24-1.35) |
| + Hypertension | 0.95 | (0.93-0.98) | 1.00 | (ref) | 1.07 | (1.01-1.13) | 0.99 | (0.89-1.10) | 1.17 | (1.12-1.22) |
| + Antihypertensive medication | 0.96 | (0.94-0.98) | 1.00 | (ref) | 1.06 | (1.00-1.12) | 1.00 | (0.90-1.10) | 1.10 | (1.06-1.15) |
| + Statin | 0.97 | (0.94-0.99) | 1.00 | (ref) | 1.06 | (1.00-1.12) | 0.99 | (0.89-1.10) | 1.11 | (1.06-1.15) |
| + Family history of CVD | 0.95 | (0.92-0.97) | 1.00 | (ref) | 1.09 | (1.03-1.15) | 1.00 | (0.90-1.11) | 1.26 | (1.21-1.31) |
| **Fully adjusted** | **0.98** | **(0.96-1.01)** | 1.00 | (ref) | **1.04** | **(0.98-1.10)** | **0.97** | **(0.88-1.08)** | **1.06** | **(1.02-1.11)** |
| **Women** |  |  |  |  |  |  |  |  |  |  |
| **Unadjusted** | **0.64** | **(0.62-0.65)** | 1.00 | (ref) | **1.67** | **(1.57-1.77)** | **1.38** | **(1.18-1.62)** | **2.19** | **(2.07-2.32)** |
| **Age-sex adjusted** | **0.86** | **(0.84-0.89)** | 1.00 | (ref) | **1.47** | **(1.38-1.56)** | **1.33** | **(1.14-1.56)** | **2.00** | **(1.89-2.12)** |
| + Ethnicity | 0.86 | (0.84-0.89) | 1.00 | (ref) | 1.48 | (1.39-1.57) | 1.34 | (1.14-1.57) | 2.01 | (1.90-2.13) |
| + IMD | 0.87 | (0.84-0.90) | 1.00 | (ref) | 1.44 | (1.36-1.53) | 1.30 | (1.11-1.52) | 1.93 | (1.82-2.04) |
| **Socio-demographic adjusted** | **0.87** | **(0.84-0.89)** | 1.00 | (ref) | **1.46** | **(1.38-1.55)** | **1.32** | **(1.12-1.54)** | **1.95** | **(1.84-2.07)** |
| + Smoking | 0.88 | (0.85-0.90) | 1.00 | (ref) | 1.44 | (1.35-1.53) | 1.29 | (1.10-1.51) | 1.95 | (1.84-2.06) |
| + Alcohol | 0.88 | (0.85-0.91) | 1.00 | (ref) | 1.43 | (1.34-1.52) | 1.27 | (1.08-1.49) | 1.86 | (1.76-1.97) |
| + Exercise | 0.87 | (0.84-0.89) | 1.00 | (ref) | 1.45 | (1.37-1.54) | 1.30 | (1.11-1.52) | 1.92 | (1.82-2.04) |
| + BMI | 0.92 | (0.89-0.95) | 1.00 | (ref) | 1.26 | (1.19-1.34) | 1.02 | (0.87-1.19) | 1.52 | (1.43-1.61) |
| + WHR | 0.91 | (0.88-0.93) | 1.00 | (ref) | 1.32 | (1.24-1.40) | 1.10 | (0.94-1.29) | 1.64 | (1.54-1.73) |
| + Processed meat consumption | 0.87 | (0.84-0.90) | 1.00 | (ref) | 1.45 | (1.37-1.54) | 1.30 | (1.11-1.53) | 1.93 | (1.83-2.05) |
| + Fruit/vegetable consumption | 0.87 | (0.84-0.90) | 1.00 | (ref) | 1.45 | (1.37-1.54) | 1.31 | (1.12-1.53) | 1.95 | (1.85-2.07) |
| **Socio-demographic and lifestyle characteristic adjusted** | **0.96** | **(0.93-0.99)** | 1.00 | (ref) | **1.18** | **(1.11-1.26)** | **0.93** | **(0.79-1.09)** | **1.40** | **(1.32-1.48)** |
| + Total cholesterol | 0.96 | (0.93-0.98) | 1.00 | (ref) | 1.18 | (1.11-1.25) | 0.92 | (0.79-1.08) | 1.29 | (1.22-1.37) |
| + eGFR | 0.96 | (0.93-0.99) | 1.00 | (ref) | 1.18 | (1.11-1.25) | 0.94 | (0.80-1.10) | 1.38 | (1.31-1.47) |
| + CRP | 0.97 | (0.94-1.00) | 1.00 | (ref) | 1.16 | (1.10-1.24) | 0.90 | (0.76-1.05) | 1.42 | (1.34-1.50) |
| + Hypertension | 0.96 | (0.93-0.99) | 1.00 | (ref) | 1.16 | (1.09-1.23) | 0.92 | (0.79-1.08) | 1.29 | (1.22-1.37) |
| + Antihypertensive medication | 0.96 | (0.94-0.99) | 1.00 | (ref) | 1.14 | (1.07-1.22) | 0.92 | (0.79-1.08) | 1.22 | (1.15-1.30) |
| + Statin | 0.97 | (0.94-1.00) | 1.00 | (ref) | 1.15 | (1.08-1.22) | 0.91 | (0.78-1.07) | 1.20 | (1.13-1.28) |
| + Family history of CVD | 0.96 | (0.93-0.99) | 1.00 | (ref) | 1.18 | (1.11-1.25) | 0.93 | (0.79-1.09) | 1.39 | (1.31-1.47) |
| **Fully adjusted** | **0.99** | **(0.96-1.02)** | 1.00 | (ref) | **1.11** | **(1.04-1.18)** | **0.88** | **(0.75-1.03)** | **1.17** | **(1.10-1.24)** |

Abbreviations: HbA1c, glycosylated haemoglobin; IMD, index of multiple deprivation; BMI, body mass index; WHR, waist-hip ratio; eGFR, estimated glomerular filtration rate; CRP, C-reactive protein; CVD, cardiovascular disease
Note: Socio-demographic adjusted model additionally includes ethnicity and IMD. Socio-demographical and lifestyle adjusted model additionally includes smoking, alcohol, exercise, BMI, WHR, processed meat consumption, and fruit/vegetable consumption. Fully adjusted model includes all variables. Individual adjustments between these four primary models are not inclusive of other adjustments made.

### Sensitivity analyses

In models additionally including angina in the definition of CAD, associations attenuated between diagnosed diabetes and CAD (HR 1.17, 95% CI 1.11-1.24 for men and HR 1.34, 95% CI 1.23-1.46 for women; p-interaction=0.0574; **eTable 3**). In contrast, associations strengthened between diagnosed diabetes and stroke after restricting the outcome to ischemic stroke only (HR 1.50, 95% CI 1.34-1.67 for men and HR 1.64, 95% CI 1.40-1.92 for women; p-interaction=0.08940; **eTable 3**). Conclusions from the primary analyses held after excluding individuals with HbA1c <20 mmol/mol (**eTable 4**), CVD events within the first 180 days (**eTable 5**), and individuals with diagnosed diabetes (**eTable 6**).

## Discussion

In this study of 427,435 adults with 51,288 incident cardiovascular events over an average 12 years of follow-up, we uncovered novel insights around sex disparities in CVD risk across the glycaemic spectrum. Absolute CVD rates were higher in men than women in all HbA1c categories. Both men and women with pre-diabetes, undiagnosed diabetes, and, more markedly, diagnosed diabetes were at higher risks of CVD than those with normal HbA1c, with relative increases in risk more pronounced in women than men. Both men and women with low-normal HbA1c had lower absolute rates of CVD than those with normal HbA1c. Lifestyle and clinical characteristics, namely obesity and use of antihypertensive medications or statins, appeared to largely account for glycaemia-associated CVD risks in both men and women, and there was little difference by sex after adjustment for these factors. Broadly, each of the six CVD outcomes showed similar patterns as the composite CVD outcome; however, men and women with diagnosed diabetes remained at elevated risk of CAD, stroke, and heart failure even after accounting for all lifestyle and clinical characteristics.

In our cohort, women with undiagnosed and diagnosed diabetes were more likely to be classified as obese than their male counterparts. While men typically get diagnosed with diabetes at a lower body mass index than women,^19^ other evidence suggests women remain more insulin sensitive despite weight gain due to their greater ability to expand subcutaneous storage capacity.^20^ In contrast, excess fat in men is stored more rapidly as ectopic fat in central and visceral depots,^5^ which in turn accelerates the development of insulin resistance.^21^ We did not observe this; more women than men in our analysis had an adverse waist-hip ratio, which is a measure strongly correlated with visceral body fat^22^ and has been demonstrated to be more strongly associated with risk of CVD in women than men.^23^ In addition, use of antihypertensive medications and statins was lower in women than men, particularly in the low-normal, normal, and pre-diabetes groups. Altogether, higher levels of obesity and lower use of treatments for CVD prevention were key factors in explaining the observed sex differences in the present study. Notably, dietary factors and clinical characteristics including total cholesterol, eGFR, C-reactive protein, or family history of CVD had relatively minimal impact in models already accounting for lifestyle characteristics, which have been suggested as potential causes in previous summary reviews.^24^

Though associations between glycaemia and the composite CVD outcome largely disappeared in fully adjusted models, we observed strong associations between diagnosed diabetes and individual CVD outcomes for men and women. Of all outcomes examined in the present study, the largest excess risks associated with diagnosed diabetes were observed for CAD, for which both men and women with diagnosed diabetes remained at ∼50-60% elevated risk in fully adjusted models. Previous meta-analyses have found larger excess risks for both men and women;^7–9^ however, many of the included studies accounted for a limited set of demographic and clinical characteristics. Our study included a wide range of characteristics, including those not typically recorded in large routinely-collected datasets. We also note that the use of antihypertensive (>60%) and lipid-lowering therapies (>70%) in people with diagnosed diabetes was high in our study cohort, which may explain why these associations were lower in our study. Conversely, we did not find any strong evidence of risk of atrial fibrillation, DVT, or PE associated with diagnosed diabetes for men or women. One potential explanation may be due to different sets of risk factors between these outcomes. In the development of a risk score based on the Framingham Heart Study data, diabetes was not a significant predictor in the 10-year risk of atrial fibrillation.^25^ With the increasing availability of imaging data, future research could further our understanding of the mechanisms between glycaemia and cardiovascular outcomes by leveraging cardiac and brain magnetic resonance imaging (MRI) data capturing preclinical stages of target organ damage.

While those with diagnosed diabetes had the highest risk, age-adjusted rates and risk of any CVD was elevated in both men and women with pre-diabetes and undiagnosed diabetes, compared to those with normal HbA1c. These risks diminished greatly or disappeared entirely in fully adjusted models; however, in outcome-specific models, men with pre-diabetes and undiagnosed diabetes remained at elevated risk of CAD. The threshold for diagnosing diabetes has been historically decided at the level of HbA1c associated with higher risk of diabetic retinopathy.^26^ Our findings are concordant with a recent systematic review and meta-analysis that demonstrated individuals with moderately elevated HbA1c below the threshold for diabetes were also at higher risk of cardiovascular outcomes.^27^ A recent meta-analysis of 51 trials demonstrated similar absolute effects and greater relative effects of initiating antihypertensive medications in patients without diabetes compared to those with diabetes.^28^ Taken together, these findings support recent recommendations for wider use of pharmaceutical strategies for the prevention of cardiovascular disease in patients without diabetes.^29^

A novel finding was some degree of protection associated with low-normal HbA1c for both men and women, which persisted after full adjustment in the CAD model. This is in contrast with previous studies that have suggested a J-shaped curve in the relationship between HbA1c and outcomes including CVD and all-cause mortality, with individuals with low-normal HbA1c at excess risk compared to normal HbA1c.^30,31^ Individuals with low-normal HbA1c may have other health conditions that place them at higher risk of adverse events. Our primary findings remained unchanged in sensitivity analyses excluding those with abnormally low HbA1c indicating chronic illness and increased mortality risk. The present study also leveraged data that allowed us to separate individuals on glucose-lowering therapies from the low-normal HbA1c group. Therefore, we are able to rule out any influence of glucose-lowering therapies on our finding of protection associated with low-normal HbA1c, highlighting the potential for non-pharmaceutical strategies to reduce risk of CVD across the glycaemic spectrum.

Our study has several key strengths, particularly a large sample size, long follow-up, highly detailed covariates with small proportions of missingness, and the availability of HbA1c on all participants, regardless of diabetes status. We also acknowledge important limitations. The overall healthier population contributing to the UK Biobank^32^ meant that we observed relatively low CVD incidence rates compared to studies in the general population. Although the underlying rates and exposure distributions may not be representative of the UK population, the associations observed between HbA1c and CVD incidence are unlikely to be biased.^33^ It is important to note our rationale of using the term “sex” rather than “gender”. For historical context on the use of “sex” and “gender” in medicine, see Marino et al^34^ and Franconi et al.^35^ In brief, “sex” refers to biological status of males and females while “gender” refers to the self-identification of an individual. Our analysis includes both lifestyle and clinical characteristics, and it is likely that biological sex and gender constructs are at play. Though the UK Biobank Data Showcase uses the term “sex” instead of “gender”, the description of this field (Data-field #31) states that “…this field may contain a mixture of the sex the NHS had recorded for the participant and self-reported sex.” Given NHS records largely include self-reported demographic information, which may capture biological status or how a person identifies, we do not have the data to appropriately distinguish between sex and gender. Current and future data collection efforts should collect these related but distinct concepts separately and include other sexes and gender identities beyond male/female or man/women. Lastly, it is possible that although we adjusted for a wide range of confounders, some residual confounding may remain. We were also unable to look at the effect of medication use in those with diagnosed diabetes as we lacked longitudinal prescription data for the full UK Biobank cohort.

## Conclusion

This is the largest study to date to investigate sex differences in the risk of CVD across the glycaemic spectrum. We demonstrated sex differences in the underlying rate and diabetes-associated risk of CVD, and showed increased risks among men and women with moderately elevated HbA1c, even below the threshold for diabetes. We extended previous evidence by showing most excess risk, and thereby differential relative risks between men and women, disappeared after accounting for lifestyle and clinical characteristics, namely measures of obesity and medications for primary prevention of CVD including antihypertensive and lipid-lowering therapies. Addressing these risk factors could reduce sex disparities in glycaemia-related risks of CVD.

## Data Availability

UK Biobank data are available to any bona fide researcher to conduct health-related research that is in the public interest at http://www.ukbiobank.ac.uk/using-the-resource/.

## Acknowledgements

This work was conducted under the approved UK Biobank project number 7661. We thank the volunteer participants of the UK Biobank, and the UK Biobank researchers.

## Ethics Approval

This study had local approval from the UK Biobank (#21893) and institutional approval from the London School of Hygiene & Tropical Medicine (#14387). All participants provided informed consent at the time of recruitment to the UK Biobank.

## Contributorship

CR, VG, LS, NC, and KB conceived the study. CR, RM, and SE curated the data. CR and KB performed the formal analysis. LS, NC, and KB acquired funding. CR and KB conducted the investigation. CR, NC, and KB designed the methodology. CR and KB managed and coordinated the project. LS and KB procured resources to carry out the study. CR and KB developed programming. LS, NC, and KB provided oversight and leadership of the project. CR and KB prepared data visualisations. CR wrote the first draft of the manuscript. All authors reviewed and edited the manuscript. CR and KB had full access to all of the data and CR had final responsibility to submit for publication. CR and KB attest that all listed authors meet authorship criteria and that no others meeting the criteria have been omitted.

