## Supplementary Appendix for "Sex-specific risks for cardiovascular disease across the glycaemic spectrum: a population-based cohort study using the UK Biobank"

### Table of Contents

**eTable 1. International Classification of Diseases 10th edition (ICD-10) codelists for cardiovascular events** **2**

**eTable 2. Sex-specific associations between HbA1c category and cardiovascular diseases** **3**

**eTable 3. Sex-specific associations amending the CAD outcome to include angina and restricting the stroke outcome to ischemic stroke only** **5**

**eTable 4. Sensitivity analysis excluding individuals with HbA1c <20 mmol/mol** **6**

**eTable 5. Sensitivity analysis excluding events occurring within 180 days of follow-up** **8**

**eTable 6. Sensitivity analysis excluding individuals with diagnosed diabetes** **10**

**eFigure 1. Schoenfeld residuals testing the proportional hazards assumption between HbA1c category and any cardiovascular disease, by sex** **12**

**The RECORD statement – checklist of items, extended from the STROBE statement, that should be reported in observational studies using routinely collected health data** **13**

**eTable 1.** International Classification of Diseases 10th edition (ICD-10) codelists for cardiovascular events

| **Outcome** | **ICD-10 codes** |
|---|---|
| Coronary artery disease (CAD) | Primary analyses included myocardial infarction (I21.*) and sudden cardiac arrest (I46.*). Sensitivity analyses additionally included angina (I20.*) |
| Atrial fibrillation (Afib) | I48.* |
| Deep vein thrombosis (DVT) | I80.1, I80.2, I80.3 |
| Pulmonary embolism (PE) | I26.* |
| Stroke | I60.*, I61.*, I62.*, I63.*, I64 |
| Heart failure (HF) | I50.* |

**eTable 2.** Sex-specific associations between HbA1c category and cardiovascular diseases

| | Unadjusted | | Age-adjusted | | + Socio-demographics | | + Lifestyle characteristics | | + Clinical characteristics | |
|---|---|---|---|---|---|---|---|---|---|---|
| | **HR** | **(95% CI)** | **HR** | **(95% CI)** | **HR** | **(95% CI)** | **HR** | **(95% CI)** | **HR** | **(95% CI)** |
| **Any cardiovascular disease*** | | | | | | | | | | |
| Men | | | | | | | | | | |
| Low-normal HbA1c | 0.72 | (0.70-0.74) | 0.86 | (0.84-0.88) | 0.87 | (0.84-0.89) | 0.95 | (0.92-0.97) | 0.98 | (0.96-1.01) |
| Normal HbA1c | 1.00 | ref | 1.00 | ref | 1.00 | ref | 1.00 | ref | 1.00 | ref |
| Pre-diabetes | 1.46 | (1.38-1.54) | 1.30 | (1.24-1.38) | 1.29 | (1.22-1.36) | 1.09 | (1.03-1.15) | 1.04 | (0.98-1.10) |
| Undiagnosed diabetes | 1.32 | (1.19-1.46) | 1.31 | (1.18-1.45) | 1.28 | (1.16-1.42) | 1.00 | (0.90-1.11) | 0.97 | (0.88-1.08) |
| Diagnosed diabetes | 1.82 | (1.75-1.89) | 1.55 | (1.49-1.61) | 1.53 | (1.47-1.59) | 1.26 | (1.21-1.32) | 1.06 | (1.02-1.11) |
| Women | | | | | | | | | | |
| Low-normal HbA1c | 0.64 | (0.62-0.65) | 0.86 | (0.84-0.89) | 0.87 | (0.84-0.89) | 0.96 | (0.93-0.99) | 0.99 | (0.96-1.02) |
| Normal HbA1c | 1.00 | ref | 1.00 | ref | 1.00 | ref | 1.00 | ref | 1.00 | ref |
| Pre-diabetes | 1.67 | (1.57-1.77) | 1.47 | (1.38-1.56) | 1.46 | (1.38-1.55) | 1.18 | (1.11-1.26) | 1.11 | (1.04-1.18) |
| Undiagnosed diabetes | 1.38 | (1.18-1.62) | 1.33 | (1.14-1.56) | 1.32 | (1.12-1.54) | 0.93 | (0.79-1.09) | 0.88 | (0.75-1.03) |
| Diagnosed diabetes | 2.19 | (2.07-2.32) | 2.00 | (1.89-2.12) | 1.95 | (1.84-2.07) | 1.40 | (1.32-1.48) | 1.17 | (1.10-1.24) |
| *p-interaction* | *<0.0001* | | *<0.0001* | | *<0.0001* | | *0.0197* | | *0.0387* | |
| **Coronary artery disease** | | | | | | | | | | |
| Men | | | | | | | | | | |
| Low-normal HbA1c | 0.64 | (0.61-0.68) | 0.74 | (0.71-0.78) | 0.75 | (0.71-0.79) | 0.85 | (0.80-0.89) | 0.90 | (0.86-0.95) |
| Normal HbA1c | 1.00 | ref | 1.00 | ref | 1.00 | ref | 1.00 | ref | 1.00 | ref |
| Pre-diabetes | 1.58 | (1.43-1.75) | 1.44 | (1.30-1.59) | 1.41 | (1.28-1.56) | 1.21 | (1.09-1.34) | 1.15 | (1.04-1.27) |
| Undiagnosed diabetes | 1.76 | (1.48-2.10) | 1.74 | (1.46-2.07) | 1.69 | (1.42-2.01) | 1.38 | (1.16-1.65) | 1.33 | (1.11-1.59) |
| Diagnosed diabetes | 2.06 | (1.92-2.21) | 1.82 | (1.69-1.95) | 1.76 | (1.64-1.89) | 1.53 | (1.42-1.65) | 1.51 | (1.39-1.63) |
| Women | | | | | | | | | | |
| Low-normal HbA1c | 0.56 | (0.52-0.60) | 0.71 | (0.65-0.76) | 0.71 | (0.66-0.77) | 0.82 | (0.76-0.89) | 0.89 | (0.82-0.96) |
| Normal HbA1c | 1.00 | ref | 1.00 | ref | 1.00 | ref | 1.00 | ref | 1.00 | ref |
| Pre-diabetes | 1.70 | (1.46-1.96) | 1.53 | (1.32-1.77) | 1.52 | (1.31-1.76) | 1.19 | (1.03-1.38) | 1.12 | (0.97-1.30) |
| Undiagnosed diabetes | 1.57 | (1.09-2.25) | 1.52 | (1.06-2.18) | 1.48 | (1.03-2.12) | 1.05 | (0.73-1.50) | 0.99 | (0.69-1.42) |
| Diagnosed diabetes | 2.76 | (2.44-3.12) | 2.55 | (2.26-2.88) | 2.46 | (2.18-2.79) | 1.79 | (1.57-2.03) | 1.80 | (1.58-2.05) |
| *p-interaction* | *<0.0001* | | *<0.0001* | | *<0.0001* | | *0.1037* | | *0.0540* | |
| **Atrial fibrillation** | | | | | | | | | | |
| Men | | | | | | | | | | |
| Low-normal HbA1c | 0.75 | (0.72-0.77) | 0.94 | (0.91-0.98) | 0.94 | (0.91-0.98) | 1.02 | (0.98-1.06) | 1.04 | (1.01-1.08) |
| Normal HbA1c | 1.00 | ref | 1.00 | ref | 1.00 | ref | 1.00 | ref | 1.00 | ref |
| Pre-diabetes | 1.50 | (1.40-1.62) | 1.30 | (1.20-1.40) | 1.31 | (1.21-1.41) | 1.09 | (1.01-1.18) | 1.03 | (0.96-1.11) |
| Undiagnosed diabetes | 1.37 | (1.18-1.58) | 1.35 | (1.17-1.56) | 1.36 | (1.18-1.57) | 1.02 | (0.88-1.18) | 1.01 | (0.87-1.17) |
| Diagnosed diabetes | 1.92 | (1.82-2.02) | 1.58 | (1.50-1.67) | 1.58 | (1.50-1.67) | 1.25 | (1.19-1.33) | 1.02 | (0.96-1.08) |
| Women | | | | | | | | | | |
| Low-normal HbA1c | 0.67 | (0.64-0.70) | 0.98 | (0.94-1.03) | 0.98 | (0.94-1.03) | 1.06 | (1.01-1.11) | 1.06 | (1.01-1.11) |
| Normal HbA1c | 1.00 | ref | 1.00 | ref | 1.00 | ref | 1.00 | ref | 1.00 | ref |
| Pre-diabetes | 1.59 | (1.45-1.74) | 1.35 | (1.23-1.48) | 1.37 | (1.24-1.50) | 1.12 | (1.02-1.24) | 1.04 | (0.95-1.15) |
| Undiagnosed diabetes | 1.29 | (1.01-1.65) | 1.22 | (0.96-1.56) | 1.25 | (0.98-1.60) | 0.90 | (0.70-1.15) | 0.85 | (0.66-1.09) |
| Diagnosed diabetes | 2.16 | (1.98-2.35) | 1.90 | (1.74-2.07) | 1.90 | (1.75-2.07) | 1.35 | (1.24-1.47) | 1.06 | (0.97-1.16) |
| *p-interaction* | *<0.0001* | | *0.0081* | | *0.0092* | | *0.3822* | | *0.6340* | |
| **Deep vein thrombosis** | | | | | | | | | | |
| Men | | | | | | | | | | |
| Low-normal HbA1c | 0.85 | (0.77-0.93) | 0.97 | (0.88-1.06) | 0.97 | (0.88-1.06) | 1.07 | (0.97-1.18) | 1.07 | (0.97-1.17) |
| Normal HbA1c | 1.00 | ref | 1.00 | ref | 1.00 | ref | 1.00 | ref | 1.00 | ref |
| Pre-diabetes | 1.22 | (0.98-1.51) | 1.11 | (0.90-1.38) | 1.10 | (0.89-1.37) | 0.92 | (0.74-1.14) | 0.92 | (0.74-1.14) |
| Undiagnosed diabetes | 1.14 | (0.76-1.73) | 1.13 | (0.75-1.71) | 1.12 | (0.74-1.69) | 0.84 | (0.55-1.27) | 0.84 | (0.56-1.27) |
| Diagnosed diabetes | 1.47 | (1.26-1.72) | 1.31 | (1.12-1.53) | 1.30 | (1.11-1.52) | 1.04 | (0.88-1.21) | 1.09 | (0.92-1.28) |
| Women | | | | | | | | | | |
| Low-normal HbA1c | 0.67 | (0.60-0.74) | 0.83 | (0.75-0.92) | 0.84 | (0.75-0.93) | 0.95 | (0.85-1.06) | 0.94 | (0.85-1.05) |
| Normal HbA1c | 1.00 | ref | 1.00 | ref | 1.00 | ref | 1.00 | ref | 1.00 | ref |
| Pre-diabetes | 1.47 | (1.18-1.83) | 1.34 | (1.08-1.66) | 1.33 | (1.07-1.65) | 1.03 | (0.83-1.29) | 1.02 | (0.82-1.27) |
| Undiagnosed diabetes | 1.52 | (0.91-2.52) | 1.47 | (0.88-2.46) | 1.46 | (0.88-2.44) | 0.98 | (0.59-1.64) | 0.98 | (0.58-1.63) |
| Diagnosed diabetes | 1.61 | (1.30-2.00) | 1.50 | (1.21-1.86) | 1.47 | (1.19-1.82) | 0.99 | (0.79-1.23) | 1.02 | (0.82-1.28) |
| *p-interaction* | *0.0013* | | *0.0408* | | *0.0451* | | *0.3596* | | *0.3659* | |
| **Pulmonary embolism** | | | | | | | | | | |
| Men | | | | | | | | | | |
| Low-normal HbA1c | 0.76 | (0.70-0.82) | 0.88 | (0.81-0.95) | 0.88 | (0.81-0.95) | 0.98 | (0.91-1.06) | 0.99 | (0.92-1.07) |
| Normal HbA1c | 1.00 | ref | 1.00 | ref | 1.00 | ref | 1.00 | ref | 1.00 | ref |
| Pre-diabetes | 1.47 | (1.25-1.72) | 1.34 | (1.14-1.57) | 1.33 | (1.13-1.56) | 1.07 | (0.91-1.26) | 1.07 | (0.91-1.25) |
| Undiagnosed diabetes | 1.33 | (0.98-1.81) | 1.32 | (0.97-1.79) | 1.30 | (0.96-1.77) | 0.94 | (0.69-1.28) | 0.93 | (0.68-1.26) |
| Diagnosed diabetes | 1.37 | (1.21-1.56) | 1.21 | (1.07-1.38) | 1.20 | (1.05-1.36) | 0.92 | (0.81-1.05) | 0.99 | (0.86-1.14) |
| Women | | | | | | | | | | |
| Low-normal HbA1c | 0.61 | (0.56-0.66) | 0.78 | (0.72-0.84) | 0.78 | (0.72-0.84) | 0.89 | (0.82-0.97) | 0.90 | (0.83-0.98) |
| Normal HbA1c | 1.00 | ref | 1.00 | ref | 1.00 | ref | 1.00 | ref | 1.00 | ref |

| | | | | | | | | | | |
|---|---|---|---|---|---|---|---|---|---|---|
| Pre-diabetes | 1.59 | (1.36-1.87) | 1.44 | (1.23-1.69) | 1.43 | (1.22-1.68) | 1.09 | (0.93-1.28) | 1.08 | (0.91-1.27) |
| Undiagnosed diabetes | 1.17 | (0.75-1.81) | 1.13 | (0.73-1.75) | 1.12 | (0.72-1.74) | 0.73 | (0.47-1.14) | 0.72 | (0.46-1.12) |
| Diagnosed diabetes | 1.64 | (1.39-1.93) | 1.51 | (1.29-1.78) | 1.48 | (1.26-1.75) | 0.98 | (0.83-1.16) | 1.03 | (0.87-1.23) |
| *p-interaction* | *0.0001* | | *0.0125* | | *0.0151* | | *0.3246* | | *0.3673* | |
| **Stroke** | | | | | | | | | | |
| Men | | | | | | | | | | |
| Low-normal HbA1c | 0.71 | (0.67-0.75) | 0.87 | (0.82-0.92) | 0.88 | (0.82-0.93) | 0.94 | (0.89-1.00) | 0.97 | (0.92-1.04) |
| Normal HbA1c | 1.00 | ref | 1.00 | ref | 1.00 | ref | 1.00 | ref | 1.00 | ref |
| Pre-diabetes | 1.44 | (1.27-1.64) | 1.26 | (1.11-1.43) | 1.23 | (1.08-1.40) | 1.10 | (0.96-1.25) | 1.04 | (0.92-1.19) |
| Undiagnosed diabetes | 1.52 | (1.21-1.91) | 1.50 | (1.19-1.88) | 1.45 | (1.15-1.82) | 1.23 | (0.98-1.55) | 1.19 | (0.95-1.51) |
| Diagnosed diabetes | 2.15 | (1.97-2.34) | 1.81 | (1.66-1.97) | 1.76 | (1.61-1.91) | 1.59 | (1.46-1.74) | 1.43 | (1.30-1.57) |
| Women | | | | | | | | | | |
| Low-normal HbA1c | 0.65 | (0.61-0.70) | 0.91 | (0.85-0.98) | 0.92 | (0.85-0.99) | 0.98 | (0.91-1.06) | 1.01 | (0.94-1.08) |
| Normal HbA1c | 1.00 | ref | 1.00 | ref | 1.00 | ref | 1.00 | ref | 1.00 | ref |
| Pre-diabetes | 1.65 | (1.44-1.91) | 1.42 | (1.24-1.64) | 1.39 | (1.20-1.60) | 1.22 | (1.06-1.41) | 1.14 | (0.99-1.32) |
| Undiagnosed diabetes | 1.33 | (0.91-1.93) | 1.26 | (0.87-1.84) | 1.22 | (0.84-1.77) | 1.01 | (0.70-1.47) | 0.95 | (0.65-1.39) |
| Diagnosed diabetes | 2.28 | (2.01-2.59) | 2.02 | (1.78-2.30) | 1.94 | (1.71-2.20) | 1.63 | (1.43-1.86) | 1.43 | (1.25-1.64) |
| *p-interaction* | *0.1116* | | *0.3654* | | *0.4287* | | *0.6118* | | *0.6596* | |
| **Heart failure** | | | | | | | | | | |
| Men | | | | | | | | | | |
| Low-normal HbA1c | 0.62 | (0.59-0.66) | 0.78 | (0.74-0.83) | 0.79 | (0.75-0.84) | 0.93 | (0.88-0.98) | 1.00 | (0.94-1.05) |
| Normal HbA1c | 1.00 | ref | 1.00 | ref | 1.00 | ref | 1.00 | ref | 1.00 | ref |
| Pre-diabetes | 1.91 | (1.73-2.10) | 1.65 | (1.50-1.82) | 1.60 | (1.45-1.76) | 1.20 | (1.09-1.33) | 1.08 | (0.98-1.19) |
| Undiagnosed diabetes | 1.99 | (1.67-2.36) | 1.96 | (1.65-2.33) | 1.86 | (1.56-2.21) | 1.21 | (1.01-1.44) | 1.15 | (0.96-1.37) |
| Diagnosed diabetes | 3.02 | (2.83-3.22) | 2.51 | (2.35-2.68) | 2.39 | (2.24-2.56) | 1.69 | (1.58-1.81) | 1.27 | (1.18-1.36) |
| Women | | | | | | | | | | |
| Low-normal HbA1c | 0.61 | (0.56-0.65) | 0.88 | (0.82-0.94) | 0.89 | (0.83-0.95) | 1.04 | (0.97-1.12) | 1.09 | (1.01-1.17) |
| Normal HbA1c | 1.00 | ref | 1.00 | ref | 1.00 | ref | 1.00 | ref | 1.00 | ref |
| Pre-diabetes | 2.43 | (2.16-2.73) | 2.07 | (1.84-2.32) | 2.00 | (1.78-2.25) | 1.43 | (1.27-1.61) | 1.25 | (1.11-1.41) |
| Undiagnosed diabetes | 2.27 | (1.71-3.01) | 2.16 | (1.62-2.86) | 2.05 | (1.54-2.73) | 1.19 | (0.89-1.58) | 1.06 | (0.79-1.40) |
| Diagnosed diabetes | 4.19 | (3.80-4.61) | 3.69 | (3.35-4.07) | 3.46 | (3.14-3.82) | 2.00 | (1.80-2.21) | 1.44 | (1.30-1.60) |
| *p-interaction* | *<0.0001* | | *<0.0001* | | *<0.0001* | | *0.0107* | | *0.0678* | |

*A composite measure of all examined outcomes

*Abbreviations:* HbA1c, glycosylated haemoglobin; HR, hazard ratio; CI, confidence interval

*Notes:* Categories were defined by baseline HbA1c levels as follows: low-normal (<35 mmol/mol or <5.5%), normal (35-41 mmol/mol or 5.5-5.9%), pre-diabetes (42-47 mmol/mol or 6.0-6.4%), undiagnosed diabetes (≥48 mmol/mol or ≥6.5%), or diagnosed diabetes defined by a diagnostic code or receipt of glucose-lowering medication. Sex-specific hazard ratios from Cox proportional hazards models, adjusted for age at study entry, socio-demographics (i.e., ethnicity, deprivation), lifestyle characteristics (i.e., smoking status, alcohol consumption, physical activity, body mass index, waist-hip ratio, processed meat and fruit and vegetable intake), and clinical characteristics (i.e., total cholesterol, estimated glomerular filtration rate, C-reactive protein, diagnosed hypertension, use of antihypertensive medication or statins, and family history of cardiovascular disease).

**eTable 3.** Sex-specific associations amending the CAD outcome to include angina and restricting the stroke outcome to ischemic stroke only

| | Unadjusted | | Age-adjusted | | + Socio-demographics | | + Lifestyle characteristics | | + Clinical characteristics | |
|---|---|---|---|---|---|---|---|---|---|---|
| | **HR** | **(95% CI)** | **HR** | **(95% CI)** | **HR** | **(95% CI)** | **HR** | **(95% CI)** | **HR** | **(95% CI)** |
| **Coronary artery disease (primary analysis)** | | | | | | | | | | |
| Men | | | | | | | | | | |
| Low-normal HbA1c | 0.64 | (0.61-0.68) | 0.74 | (0.71-0.78) | 0.75 | (0.71-0.79) | 0.85 | (0.80-0.89) | 0.90 | (0.86-0.95) |
| Normal HbA1c | 1.00 | ref | 1.00 | ref | 1.00 | ref | 1.00 | ref | 1.00 | ref |
| Pre-diabetes | 1.58 | (1.43-1.75) | 1.44 | (1.30-1.59) | 1.41 | (1.28-1.56) | 1.21 | (1.09-1.34) | 1.15 | (1.04-1.27) |
| Undiagnosed diabetes | 1.76 | (1.48-2.10) | 1.74 | (1.46-2.07) | 1.69 | (1.42-2.01) | 1.38 | (1.16-1.65) | 1.33 | (1.11-1.59) |
| Diagnosed diabetes | 2.06 | (1.92-2.21) | 1.82 | (1.69-1.95) | 1.76 | (1.64-1.89) | 1.53 | (1.42-1.65) | 1.51 | (1.39-1.63) |
| Women | | | | | | | | | | |
| Low-normal HbA1c | 0.56 | (0.52-0.60) | 0.71 | (0.65-0.76) | 0.71 | (0.66-0.77) | 0.82 | (0.76-0.89) | 0.89 | (0.82-0.96) |
| Normal HbA1c | 1.00 | ref | 1.00 | ref | 1.00 | ref | 1.00 | ref | 1.00 | ref |
| Pre-diabetes | 1.70 | (1.46-1.96) | 1.53 | (1.32-1.77) | 1.52 | (1.31-1.76) | 1.19 | (1.03-1.38) | 1.12 | (0.97-1.30) |
| Undiagnosed diabetes | 1.57 | (1.09-2.25) | 1.52 | (1.06-2.18) | 1.48 | (1.03-2.12) | 1.05 | (0.73-1.50) | 0.99 | (0.69-1.42) |
| Diagnosed diabetes | 2.76 | (2.44-3.12) | 2.55 | (2.26-2.88) | 2.46 | (2.18-2.79) | 1.79 | (1.57-2.03) | 1.80 | (1.58-2.05) |
| *p-interaction* | *<0.0001* | | *<0.0001* | | *<0.0001* | | *0.1037* | | *0.0540* | |
| **Coronary artery disease including angina (sensitivity analysis)** | | | | | | | | | | |
| Men | | | | | | | | | | |
| Low-normal HbA1c | 0.65 | (0.62-0.67) | 0.76 | (0.73-0.78) | 0.76 | (0.74-0.79) | 0.85 | (0.81-0.88) | 0.91 | (0.88-0.95) |
| Normal HbA1c | 1.00 | ref | 1.00 | ref | 1.00 | ref | 1.00 | ref | 1.00 | ref |
| Pre-diabetes | 1.55 | (1.44-1.68) | 1.41 | (1.31-1.52) | 1.38 | (1.28-1.49) | 1.17 | (1.08-1.26) | 1.09 | (1.01-1.17) |
| Undiagnosed diabetes | 1.42 | (1.23-1.64) | 1.41 | (1.22-1.63) | 1.36 | (1.18-1.57) | 1.09 | (0.94-1.26) | 1.06 | (0.91-1.22) |
| Diagnosed diabetes | 2.10 | (1.99-2.21) | 1.83 | (1.74-1.93) | 1.77 | (1.68-1.87) | 1.48 | (1.40-1.57) | 1.17 | (1.11-1.24) |
| Women | | | | | | | | | | |
| Low-normal HbA1c | 0.56 | (0.53-0.59) | 0.73 | (0.69-0.77) | 0.73 | (0.70-0.77) | 0.84 | (0.80-0.88) | 0.90 | (0.86-0.95) |
| Normal HbA1c | 1.00 | ref | 1.00 | ref | 1.00 | ref | 1.00 | ref | 1.00 | ref |
| Pre-diabetes | 1.82 | (1.66-2.00) | 1.64 | (1.49-1.80) | 1.61 | (1.47-1.77) | 1.27 | (1.16-1.39) | 1.16 | (1.06-1.27) |
| Undiagnosed diabetes | 1.70 | (1.36-2.13) | 1.64 | (1.31-2.06) | 1.58 | (1.26-1.98) | 1.08 | (0.86-1.36) | 1.00 | (0.80-1.26) |
| Diagnosed diabetes | 2.73 | (2.52-2.96) | 2.52 | (2.32-2.73) | 2.42 | (2.23-2.62) | 1.69 | (1.55-1.84) | 1.34 | (1.23-1.46) |
| *p-interaction* | *<0.0001* | | *<0.0001* | | *<0.0001* | | *0.0671* | | *0.0574* | |
| **Any stroke (primary analysis)** | | | | | | | | | | |
| Men | | | | | | | | | | |
| Low-normal HbA1c | 0.71 | (0.67-0.75) | 0.87 | (0.82-0.92) | 0.88 | (0.82-0.93) | 0.94 | (0.89-1.00) | 0.97 | (0.92-1.04) |
| Normal HbA1c | 1.00 | ref | 1.00 | ref | 1.00 | ref | 1.00 | ref | 1.00 | ref |
| Pre-diabetes | 1.44 | (1.27-1.64) | 1.26 | (1.11-1.43) | 1.23 | (1.08-1.40) | 1.10 | (0.96-1.25) | 1.04 | (0.92-1.19) |
| Undiagnosed diabetes | 1.52 | (1.21-1.91) | 1.50 | (1.19-1.88) | 1.45 | (1.15-1.82) | 1.23 | (0.98-1.55) | 1.19 | (0.95-1.51) |
| Diagnosed diabetes | 2.15 | (1.97-2.34) | 1.81 | (1.66-1.97) | 1.76 | (1.61-1.91) | 1.59 | (1.46-1.74) | 1.43 | (1.30-1.57) |
| Women | | | | | | | | | | |
| Low-normal HbA1c | 0.65 | (0.61-0.70) | 0.91 | (0.85-0.98) | 0.92 | (0.85-0.99) | 0.98 | (0.91-1.06) | 1.01 | (0.94-1.08) |
| Normal HbA1c | 1.00 | ref | 1.00 | ref | 1.00 | ref | 1.00 | ref | 1.00 | ref |
| Pre-diabetes | 1.65 | (1.44-1.91) | 1.42 | (1.24-1.64) | 1.39 | (1.20-1.60) | 1.22 | (1.06-1.41) | 1.14 | (0.99-1.32) |
| Undiagnosed diabetes | 1.33 | (0.91-1.93) | 1.26 | (0.87-1.84) | 1.22 | (0.84-1.77) | 1.01 | (0.70-1.47) | 0.95 | (0.65-1.39) |
| Diagnosed diabetes | 2.28 | (2.01-2.59) | 2.02 | (1.78-2.30) | 1.94 | (1.71-2.20) | 1.63 | (1.43-1.86) | 1.43 | (1.25-1.64) |
| *p-interaction* | *0.1116* | | *0.3654* | | *0.4287* | | *0.6118* | | *0.6596* | |
| **Ischemic stroke (sensitivity analysis)** | | | | | | | | | | |
| Men | | | | | | | | | | |
| Low-normal HbA1c | 0.68 | (0.63-0.73) | 0.83 | (0.78-0.90) | 0.84 | (0.78-0.91) | 0.92 | (0.85-0.99) | 0.96 | (0.89-1.03) |
| Normal HbA1c | 1.00 | ref | 1.00 | ref | 1.00 | ref | 1.00 | ref | 1.00 | ref |
| Pre-diabetes | 1.49 | (1.28-1.73) | 1.30 | (1.12-1.51) | 1.26 | (1.09-1.47) | 1.10 | (0.95-1.28) | 1.05 | (0.90-1.22) |
| Undiagnosed diabetes | 1.65 | (1.27-2.15) | 1.62 | (1.25-2.11) | 1.56 | (1.20-2.03) | 1.29 | (0.99-1.68) | 1.25 | (0.96-1.62) |
| Diagnosed diabetes | 2.29 | (2.07-2.52) | 1.91 | (1.73-2.11) | 1.85 | (1.68-2.05) | 1.64 | (1.48-1.82) | 1.50 | (1.34-1.67) |
| Women | | | | | | | | | | |
| Low-normal HbA1c | 0.61 | (0.56-0.67) | 0.86 | (0.78-0.94) | 0.87 | (0.79-0.95) | 0.94 | (0.86-1.04) | 0.97 | (0.89-1.07) |
| Normal HbA1c | 1.00 | ref | 1.00 | ref | 1.00 | ref | 1.00 | ref | 1.00 | ref |
| Pre-diabetes | 1.68 | (1.41-2.01) | 1.44 | (1.21-1.72) | 1.40 | (1.17-1.66) | 1.20 | (1.00-1.43) | 1.11 | (0.93-1.33) |
| Undiagnosed diabetes | 1.87 | (1.26-2.77) | 1.77 | (1.19-2.63) | 1.69 | (1.14-2.52) | 1.35 | (0.91-2.01) | 1.26 | (0.85-1.88) |
| Diagnosed diabetes | 2.70 | (2.33-3.13) | 2.38 | (2.05-2.76) | 2.27 | (1.95-2.63) | 1.84 | (1.58-2.15) | 1.64 | (1.40-1.92) |
| *p-interaction* | *0.0364* | | *0.1971* | | *0.2723* | | *0.7787* | | *0.8940* | |

*Abbreviations:* HbA1c, glycosylated haemoglobin; HR, hazard ratio; CI, confidence interval

*Notes:* Categories were defined by baseline HbA1c levels as follows: low-normal (<35 mmol/mol or <5.5%), normal (35-41 mmol/mol or 5.5-5.9%), pre-diabetes (42-47 mmol/mol or 6.0-6.4%), undiagnosed diabetes (≥48 mmol/mol or ≥6.5%), or diagnosed diabetes defined by a diagnostic code or receipt of glucose-lowering medication. In primary analyses, coronary artery disease included myocardial infarction and sudden cardiac arrest. This table additionally includes angina in the outcome definition.

**eTable 4.** Sensitivity analysis excluding individuals with HbA1c <20 mmol/mol

| | Unadjusted | | Age-adjusted | | + Socio-demographics | | + Lifestyle characteristics | | + Clinical characteristics | |
|---|---|---|---|---|---|---|---|---|---|---|
| | **HR** | **(95% CI)** | **HR** | **(95% CI)** | **HR** | **(95% CI)** | **HR** | **(95% CI)** | **HR** | **(95% CI)** |
| **Any cardiovascular disease*** | | | | | | | | | | |
| Men | | | | | | | | | | |
| Low-normal HbA1c | 0.72 | (0.70-0.74) | 0.86 | (0.84-0.88) | 0.87 | (0.84-0.89) | 0.95 | (0.92-0.97) | 0.98 | (0.96-1.01) |
| Normal HbA1c | 1.00 | ref | 1.00 | ref | 1.00 | ref | 1.00 | ref | 1.00 | ref |
| Pre-diabetes | 1.46 | (1.38-1.54) | 1.30 | (1.24-1.38) | 1.29 | (1.22-1.36) | 1.09 | (1.03-1.15) | 1.04 | (0.98-1.10) |
| Undiagnosed diabetes | 1.32 | (1.19-1.46) | 1.31 | (1.18-1.45) | 1.28 | (1.16-1.42) | 1.00 | (0.90-1.11) | 0.97 | (0.88-1.08) |
| Diagnosed diabetes | 1.82 | (1.75-1.89) | 1.55 | (1.49-1.61) | 1.53 | (1.47-1.59) | 1.26 | (1.21-1.31) | 1.06 | (1.02-1.11) |
| Women | | | | | | | | | | |
| Low-normal HbA1c | 0.64 | (0.62-0.65) | 0.86 | (0.84-0.89) | 0.87 | (0.84-0.89) | 0.96 | (0.93-0.99) | 0.98 | (0.96-1.02) |
| Normal HbA1c | 1.00 | ref | 1.00 | ref | 1.00 | ref | 1.00 | ref | 1.00 | ref |
| Pre-diabetes | 1.67 | (1.57-1.77) | 1.47 | (1.38-1.56) | 1.46 | (1.38-1.55) | 1.18 | (1.11-1.26) | 1.11 | (1.04-1.18) |
| Undiagnosed diabetes | 1.38 | (1.18-1.62) | 1.33 | (1.14-1.56) | 1.32 | (1.12-1.54) | 0.93 | (0.79-1.09) | 0.88 | (0.75-1.03) |
| Diagnosed diabetes | 2.19 | (2.07-2.32) | 2.00 | (1.89-2.12) | 1.95 | (1.84-2.07) | 1.40 | (1.32-1.48) | 1.17 | (1.10-1.24) |
| *p-interaction* | *<0.0001* | | *<0.0001* | | *<0.0001* | | *0.0197* | | *0.0379* | |
| **Coronary artery disease** | | | | | | | | | | |
| Men | | | | | | | | | | |
| Low-normal HbA1c | 0.64 | (0.61-0.68) | 0.75 | (0.71-0.78) | 0.75 | (0.71-0.79) | 0.85 | (0.80-0.89) | 0.90 | (0.86-0.95) |
| Normal HbA1c | 1.00 | ref | 1.00 | ref | 1.00 | ref | 1.00 | ref | 1.00 | ref |
| Pre-diabetes | 1.58 | (1.43-1.75) | 1.44 | (1.30-1.59) | 1.41 | (1.28-1.56) | 1.21 | (1.09-1.34) | 1.15 | (1.04-1.27) |
| Undiagnosed diabetes | 1.76 | (1.48-2.10) | 1.74 | (1.46-2.07) | 1.69 | (1.42-2.01) | 1.38 | (1.16-1.65) | 1.33 | (1.11-1.59) |
| Diagnosed diabetes | 2.06 | (1.92-2.21) | 1.82 | (1.69-1.95) | 1.76 | (1.64-1.89) | 1.53 | (1.42-1.65) | 1.51 | (1.39-1.63) |
| Women | | | | | | | | | | |
| Low-normal HbA1c | 0.56 | (0.52-0.60) | 0.71 | (0.65-0.77) | 0.71 | (0.66-0.77) | 0.82 | (0.76-0.89) | 0.89 | (0.82-0.96) |
| Normal HbA1c | 1.00 | ref | 1.00 | ref | 1.00 | ref | 1.00 | ref | 1.00 | ref |
| Pre-diabetes | 1.70 | (1.46-1.96) | 1.53 | (1.32-1.77) | 1.52 | (1.31-1.76) | 1.19 | (1.03-1.38) | 1.12 | (0.97-1.30) |
| Undiagnosed diabetes | 1.57 | (1.09-2.25) | 1.52 | (1.06-2.18) | 1.48 | (1.03-2.12) | 1.05 | (0.73-1.50) | 0.99 | (0.69-1.42) |
| Diagnosed diabetes | 2.76 | (2.44-3.12) | 2.55 | (2.26-2.88) | 2.46 | (2.18-2.79) | 1.79 | (1.57-2.03) | 1.80 | (1.58-2.05) |
| *p-interaction* | *<0.0001* | | *<0.0001* | | *<0.0001* | | *0.1054* | | *0.0548* | |
| **Atrial fibrillation** | | | | | | | | | | |
| Men | | | | | | | | | | |
| Low-normal HbA1c | 0.75 | (0.72-0.77) | 0.94 | (0.91-0.98) | 0.94 | (0.91-0.98) | 1.02 | (0.98-1.06) | 1.04 | (1.01-1.08) |
| Normal HbA1c | 1.00 | ref | 1.00 | ref | 1.00 | ref | 1.00 | ref | 1.00 | ref |
| Pre-diabetes | 1.50 | (1.40-1.62) | 1.30 | (1.20-1.40) | 1.31 | (1.21-1.41) | 1.09 | (1.01-1.18) | 1.03 | (0.96-1.11) |
| Undiagnosed diabetes | 1.37 | (1.18-1.58) | 1.35 | (1.17-1.56) | 1.36 | (1.18-1.57) | 1.02 | (0.88-1.18) | 1.01 | (0.87-1.17) |
| Diagnosed diabetes | 1.92 | (1.82-2.02) | 1.58 | (1.50-1.67) | 1.58 | (1.50-1.67) | 1.25 | (1.19-1.33) | 1.02 | (0.96-1.08) |
| Women | | | | | | | | | | |
| Low-normal HbA1c | 0.67 | (0.64-0.70) | 0.98 | (0.94-1.03) | 0.98 | (0.94-1.03) | 1.06 | (1.01-1.11) | 1.06 | (1.01-1.11) |
| Normal HbA1c | 1.00 | ref | 1.00 | ref | 1.00 | ref | 1.00 | ref | 1.00 | ref |
| Pre-diabetes | 1.59 | (1.45-1.74) | 1.35 | (1.23-1.48) | 1.37 | (1.24-1.50) | 1.12 | (1.02-1.24) | 1.04 | (0.95-1.15) |
| Undiagnosed diabetes | 1.29 | (1.01-1.65) | 1.22 | (0.96-1.56) | 1.25 | (0.98-1.60) | 0.90 | (0.70-1.15) | 0.85 | (0.66-1.09) |
| Diagnosed diabetes | 2.16 | (1.98-2.35) | 1.90 | (1.74-2.07) | 1.90 | (1.75-2.07) | 1.35 | (1.24-1.47) | 1.06 | (0.97-1.16) |
| *p-interaction* | *<0.0001* | | *0.0081* | | *0.0092* | | *0.3787* | | *0.6329* | |
| **Deep vein thrombosis** | | | | | | | | | | |
| Men | | | | | | | | | | |
| Low-normal HbA1c | 0.84 | (0.77-0.92) | 0.96 | (0.88-1.06) | 0.97 | (0.88-1.06) | 1.07 | (0.97-1.17) | 1.06 | (0.97-1.17) |
| Normal HbA1c | 1.00 | ref | 1.00 | ref | 1.00 | ref | 1.00 | ref | 1.00 | ref |
| Pre-diabetes | 1.22 | (0.98-1.51) | 1.11 | (0.90-1.38) | 1.10 | (0.89-1.37) | 0.92 | (0.74-1.14) | 0.92 | (0.74-1.14) |
| Undiagnosed diabetes | 1.14 | (0.76-1.73) | 1.13 | (0.75-1.71) | 1.12 | (0.74-1.69) | 0.84 | (0.55-1.27) | 0.84 | (0.56-1.28) |
| Diagnosed diabetes | 1.47 | (1.26-1.72) | 1.31 | (1.12-1.53) | 1.30 | (1.12-1.52) | 1.04 | (0.88-1.22) | 1.09 | (0.92-1.28) |
| Women | | | | | | | | | | |
| Low-normal HbA1c | 0.67 | (0.60-0.74) | 0.83 | (0.75-0.93) | 0.84 | (0.75-0.93) | 0.95 | (0.86-1.06) | 0.95 | (0.85-1.05) |
| Normal HbA1c | 1.00 | ref | 1.00 | ref | 1.00 | ref | 1.00 | ref | 1.00 | ref |
| Pre-diabetes | 1.47 | (1.18-1.83) | 1.34 | (1.08-1.67) | 1.33 | (1.07-1.65) | 1.03 | (0.83-1.29) | 1.02 | (0.82-1.28) |
| Undiagnosed diabetes | 1.52 | (0.91-2.52) | 1.47 | (0.88-2.46) | 1.46 | (0.88-2.44) | 0.98 | (0.59-1.64) | 0.98 | (0.58-1.63) |
| Diagnosed diabetes | 1.61 | (1.30-2.00) | 1.50 | (1.21-1.86) | 1.47 | (1.19-1.83) | 0.99 | (0.79-1.23) | 1.02 | (0.82-1.28) |
| *p-interaction* | *0.0017* | | *0.0482* | | *0.0532* | | *0.3951* | | *0.4011* | |
| **Pulmonary embolism** | | | | | | | | | | |
| Men | | | | | | | | | | |
| Low-normal HbA1c | 0.76 | (0.70-0.82) | 0.88 | (0.81-0.95) | 0.88 | (0.82-0.95) | 0.98 | (0.91-1.06) | 0.99 | (0.92-1.07) |
| Normal HbA1c | 1.00 | ref | 1.00 | ref | 1.00 | ref | 1.00 | ref | 1.00 | ref |
| Pre-diabetes | 1.47 | (1.25-1.72) | 1.34 | (1.14-1.57) | 1.33 | (1.13-1.56) | 1.07 | (0.91-1.26) | 1.07 | (0.91-1.25) |
| Undiagnosed diabetes | 1.33 | (0.98-1.81) | 1.32 | (0.97-1.79) | 1.30 | (0.96-1.77) | 0.94 | (0.69-1.28) | 0.93 | (0.68-1.26) |
| Diagnosed diabetes | 1.37 | (1.21-1.56) | 1.21 | (1.07-1.38) | 1.20 | (1.05-1.36) | 0.92 | (0.81-1.05) | 0.99 | (0.87-1.14) |
| Women | | | | | | | | | | |
| Low-normal HbA1c | 0.61 | (0.56-0.66) | 0.78 | (0.71-0.84) | 0.78 | (0.72-0.84) | 0.89 | (0.82-0.97) | 0.90 | (0.83-0.98) |
| Normal HbA1c | 1.00 | ref | 1.00 | ref | 1.00 | ref | 1.00 | ref | 1.00 | ref |

| | | | | | | | | | | |
|---|---|---|---|---|---|---|---|---|---|---|
| Pre-diabetes | 1.59 | (1.36-1.87) | 1.44 | (1.23-1.69) | 1.43 | (1.22-1.68) | 1.09 | (0.93-1.29) | 1.08 | (0.92-1.27) |
| Undiagnosed diabetes | 1.17 | (0.75-1.81) | 1.13 | (0.73-1.75) | 1.12 | (0.72-1.74) | 0.73 | (0.47-1.14) | 0.72 | (0.46-1.12) |
| Diagnosed diabetes | 1.64 | (1.39-1.93) | 1.51 | (1.29-1.78) | 1.48 | (1.26-1.75) | 0.98 | (0.83-1.16) | 1.03 | (0.87-1.23) |
| *p-interaction* | *0.0001* | | *0.0115* | | *0.0139* | | *0.3094* | | *0.3507* | |
| **Stroke** | | | | | | | | | | |
| Men | | | | | | | | | | |
| Low-normal HbA1c | 0.71 | (0.67-0.75) | 0.87 | (0.82-0.92) | 0.88 | (0.82-0.93) | 0.94 | (0.89-1.00) | 0.98 | (0.92-1.04) |
| Normal HbA1c | 1.00 | ref | 1.00 | ref | 1.00 | ref | 1.00 | ref | 1.00 | ref |
| Pre-diabetes | 1.44 | (1.27-1.64) | 1.26 | (1.11-1.43) | 1.23 | (1.08-1.40) | 1.10 | (0.96-1.25) | 1.04 | (0.92-1.19) |
| Undiagnosed diabetes | 1.52 | (1.21-1.91) | 1.50 | (1.19-1.88) | 1.45 | (1.15-1.82) | 1.23 | (0.98-1.55) | 1.20 | (0.95-1.51) |
| Diagnosed diabetes | 2.15 | (1.97-2.34) | 1.81 | (1.66-1.97) | 1.76 | (1.61-1.92) | 1.59 | (1.46-1.74) | 1.43 | (1.30-1.57) |
| Women | | | | | | | | | | |
| Low-normal HbA1c | 0.65 | (0.61-0.70) | 0.91 | (0.84-0.98) | 0.92 | (0.85-0.98) | 0.98 | (0.91-1.06) | 1.00 | (0.93-1.08) |
| Normal HbA1c | 1.00 | ref | 1.00 | ref | 1.00 | ref | 1.00 | ref | 1.00 | ref |
| Pre-diabetes | 1.65 | (1.44-1.91) | 1.43 | (1.24-1.64) | 1.39 | (1.20-1.60) | 1.22 | (1.06-1.41) | 1.14 | (0.99-1.32) |
| Undiagnosed diabetes | 1.33 | (0.91-1.93) | 1.26 | (0.87-1.84) | 1.22 | (0.84-1.77) | 1.01 | (0.70-1.47) | 0.95 | (0.65-1.39) |
| Diagnosed diabetes | 2.28 | (2.01-2.59) | 2.02 | (1.78-2.30) | 1.94 | (1.71-2.20) | 1.63 | (1.43-1.86) | 1.43 | (1.25-1.64) |
| *p-interaction* | *0.0990* | | *0.3739* | | *0.4397* | | *0.6282* | | *0.6730* | |
| **Heart failure** | | | | | | | | | | |
| Men | | | | | | | | | | |
| Low-normal HbA1c | 0.62 | (0.59-0.66) | 0.78 | (0.74-0.83) | 0.79 | (0.75-0.84) | 0.93 | (0.88-0.98) | 1.00 | (0.94-1.05) |
| Normal HbA1c | 1.00 | ref | 1.00 | ref | 1.00 | ref | 1.00 | ref | 1.00 | ref |
| Pre-diabetes | 1.91 | (1.73-2.10) | 1.65 | (1.50-1.82) | 1.60 | (1.45-1.76) | 1.20 | (1.09-1.33) | 1.08 | (0.98-1.19) |
| Undiagnosed diabetes | 1.99 | (1.67-2.36) | 1.96 | (1.65-2.33) | 1.86 | (1.56-2.21) | 1.21 | (1.01-1.44) | 1.15 | (0.96-1.37) |
| Diagnosed diabetes | 3.02 | (2.83-3.22) | 2.51 | (2.35-2.68) | 2.40 | (2.24-2.56) | 1.69 | (1.58-1.81) | 1.27 | (1.18-1.36) |
| Women | | | | | | | | | | |
| Low-normal HbA1c | 0.60 | (0.56-0.65) | 0.88 | (0.82-0.94) | 0.89 | (0.83-0.95) | 1.04 | (0.97-1.12) | 1.09 | (1.01-1.17) |
| Normal HbA1c | 1.00 | ref | 1.00 | ref | 1.00 | ref | 1.00 | ref | 1.00 | ref |
| Pre-diabetes | 2.43 | (2.16-2.73) | 2.07 | (1.84-2.32) | 2.00 | (1.78-2.25) | 1.43 | (1.27-1.61) | 1.25 | (1.11-1.41) |
| Undiagnosed diabetes | 2.27 | (1.71-3.01) | 2.16 | (1.62-2.86) | 2.05 | (1.55-2.73) | 1.19 | (0.89-1.58) | 1.05 | (0.79-1.40) |
| Diagnosed diabetes | 4.19 | (3.80-4.61) | 3.69 | (3.35-4.07) | 3.46 | (3.14-3.82) | 1.99 | (1.80-2.21) | 1.44 | (1.30-1.60) |
| *p-interaction* | *<0.0001* | | *<0.0001* | | *<0.0001* | | *0.0114* | | *0.0712* | |

*A composite measure of all examined outcomes

*Abbreviations:* HbA1c, glycosylated haemoglobin; HR, hazard ratio; CI, confidence interval

*Notes:* Categories were defined by baseline HbA1c levels as follows: low-normal (<35 mmol/mol or <5.5%), normal (35-41 mmol/mol or 5.5-5.9%), pre-diabetes (42-47 mmol/mol or 6.0-6.4%), undiagnosed diabetes (≥48 mmol/mol or ≥6.5%), or diagnosed diabetes defined by a diagnostic code or receipt of glucose-lowering medication. Sex-specific hazard ratios from Cox proportional hazards models, adjusted for age at study entry, socio-demographics (i.e., ethnicity, deprivation), lifestyle characteristics (i.e., smoking status, alcohol consumption, physical activity, body mass index, waist-hip ratio, processed meat and fruit and vegetable intake), and clinical characteristics (i.e., total cholesterol, estimated glomerular filtration rate, C-reactive protein, diagnosed hypertension, use of antihypertensive medication or statins, and family history of cardiovascular disease).

**eTable 5.** Sensitivity analysis excluding events occurring within 180 days of follow-up

| | Unadjusted | | Age-adjusted | | + Socio-demographics | | + Lifestyle characteristics | | + Clinical characteristics | |
|---|---|---|---|---|---|---|---|---|---|---|
| | **HR** | **(95% CI)** | **HR** | **(95% CI)** | **HR** | **(95% CI)** | **HR** | **(95% CI)** | **HR** | **(95% CI)** |
| **Any cardiovascular disease*** | | | | | | | | | | |
| Men | | | | | | | | | | |
| Low-normal HbA1c | 0.72 | (0.70-0.74) | 0.86 | (0.84-0.88) | 0.87 | (0.84-0.89) | 0.95 | (0.93-0.97) | 0.98 | (0.96-1.01) |
| Normal HbA1c | 1.00 | ref | 1.00 | ref | 1.00 | ref | 1.00 | ref | 1.00 | ref |
| Pre-diabetes | 1.46 | (1.38-1.54) | 1.30 | (1.23-1.37) | 1.29 | (1.22-1.36) | 1.08 | (1.02-1.14) | 1.04 | (0.98-1.09) |
| Undiagnosed diabetes | 1.31 | (1.18-1.46) | 1.30 | (1.17-1.44) | 1.28 | (1.15-1.42) | 0.99 | (0.89-1.10) | 0.97 | (0.87-1.07) |
| Diagnosed diabetes | 1.81 | (1.74-1.88) | 1.54 | (1.48-1.61) | 1.52 | (1.46-1.58) | 1.26 | (1.20-1.31) | 1.06 | (1.02-1.11) |
| Women | | | | | | | | | | |
| Low-normal HbA1c | 0.64 | (0.62-0.65) | 0.86 | (0.84-0.89) | 0.87 | (0.84-0.89) | 0.96 | (0.93-0.99) | 0.98 | (0.95-1.01) |
| Normal HbA1c | 1.00 | ref | 1.00 | ref | 1.00 | ref | 1.00 | ref | 1.00 | ref |
| Pre-diabetes | 1.63 | (1.53-1.73) | 1.43 | (1.35-1.53) | 1.43 | (1.34-1.52) | 1.15 | (1.08-1.23) | 1.08 | (1.02-1.15) |
| Undiagnosed diabetes | 1.33 | (1.13-1.57) | 1.28 | (1.09-1.51) | 1.27 | (1.08-1.49) | 0.90 | (0.76-1.06) | 0.85 | (0.73-1.00) |
| Diagnosed diabetes | 2.18 | (2.06-2.31) | 1.99 | (1.88-2.11) | 1.94 | (1.83-2.06) | 1.39 | (1.31-1.47) | 1.17 | (1.10-1.24) |
| *p-interaction* | *<0.0001* | | *<0.0001* | | *<0.0001* | | *0.0315* | | *0.0464* | |
| **Coronary artery disease** | | | | | | | | | | |
| Men | | | | | | | | | | |
| Low-normal HbA1c | 0.65 | (0.61-0.68) | 0.75 | (0.71-0.79) | 0.75 | (0.71-0.79) | 0.85 | (0.80-0.89) | 0.90 | (0.86-0.95) |
| Normal HbA1c | 1.00 | ref | 1.00 | ref | 1.00 | ref | 1.00 | ref | 1.00 | ref |
| Pre-diabetes | 1.58 | (1.43-1.75) | 1.44 | (1.30-1.59) | 1.41 | (1.28-1.57) | 1.20 | (1.09-1.33) | 1.15 | (1.03-1.27) |
| Undiagnosed diabetes | 1.76 | (1.47-2.10) | 1.73 | (1.45-2.07) | 1.69 | (1.41-2.01) | 1.37 | (1.15-1.64) | 1.32 | (1.11-1.58) |
| Diagnosed diabetes | 2.09 | (1.94-2.24) | 1.84 | (1.71-1.98) | 1.78 | (1.66-1.91) | 1.54 | (1.43-1.66) | 1.51 | (1.40-1.64) |
| Women | | | | | | | | | | |
| Low-normal HbA1c | 0.56 | (0.52-0.61) | 0.71 | (0.66-0.77) | 0.72 | (0.66-0.77) | 0.83 | (0.76-0.90) | 0.89 | (0.82-0.97) |
| Normal HbA1c | 1.00 | ref | 1.00 | ref | 1.00 | ref | 1.00 | ref | 1.00 | ref |
| Pre-diabetes | 1.69 | (1.45-1.96) | 1.52 | (1.31-1.77) | 1.51 | (1.30-1.75) | 1.19 | (1.02-1.38) | 1.12 | (0.96-1.30) |
| Undiagnosed diabetes | 1.55 | (1.07-2.24) | 1.50 | (1.04-2.16) | 1.45 | (1.01-2.10) | 1.03 | (0.71-1.49) | 0.97 | (0.67-1.41) |
| Diagnosed diabetes | 2.76 | (2.44-3.12) | 2.55 | (2.26-2.89) | 2.46 | (2.18-2.79) | 1.78 | (1.57-2.03) | 1.79 | (1.57-2.04) |
| *p-interaction* | *<0.0001* | | *0.0001* | | *0.0001* | | *0.1394* | | *0.0763* | |
| **Atrial fibrillation** | | | | | | | | | | |
| Men | | | | | | | | | | |
| Low-normal HbA1c | 0.75 | (0.72-0.77) | 0.94 | (0.91-0.98) | 0.94 | (0.91-0.98) | 1.02 | (0.98-1.06) | 1.04 | (1.01-1.08) |
| Normal HbA1c | 1.00 | ref | 1.00 | ref | 1.00 | ref | 1.00 | ref | 1.00 | ref |
| Pre-diabetes | 1.50 | (1.39-1.62) | 1.29 | (1.20-1.40) | 1.30 | (1.21-1.41) | 1.09 | (1.01-1.18) | 1.03 | (0.95-1.11) |
| Undiagnosed diabetes | 1.36 | (1.17-1.57) | 1.34 | (1.15-1.55) | 1.35 | (1.17-1.56) | 1.01 | (0.87-1.17) | 1.00 | (0.86-1.16) |
| Diagnosed diabetes | 1.92 | (1.82-2.03) | 1.58 | (1.50-1.67) | 1.59 | (1.50-1.68) | 1.26 | (1.19-1.33) | 1.02 | (0.96-1.08) |
| Women | | | | | | | | | | |
| Low-normal HbA1c | 0.66 | (0.63-0.70) | 0.98 | (0.94-1.03) | 0.98 | (0.93-1.02) | 1.06 | (1.01-1.11) | 1.06 | (1.01-1.11) |
| Normal HbA1c | 1.00 | ref | 1.00 | ref | 1.00 | ref | 1.00 | ref | 1.00 | ref |
| Pre-diabetes | 1.55 | (1.41-1.71) | 1.32 | (1.20-1.45) | 1.34 | (1.21-1.47) | 1.10 | (0.99-1.21) | 1.02 | (0.92-1.12) |
| Undiagnosed diabetes | 1.25 | (0.97-1.61) | 1.19 | (0.92-1.53) | 1.21 | (0.94-1.56) | 0.87 | (0.67-1.12) | 0.82 | (0.64-1.06) |
| Diagnosed diabetes | 2.14 | (1.97-2.34) | 1.89 | (1.73-2.06) | 1.89 | (1.74-2.06) | 1.34 | (1.23-1.46) | 1.06 | (0.96-1.16) |
| *p-interaction* | *<0.0001* | | *0.0131* | | *0.0155* | | *0.4301* | | *0.6141* | |
| **Deep vein thrombosis** | | | | | | | | | | |
| Men | | | | | | | | | | |
| Low-normal HbA1c | 0.84 | (0.77-0.93) | 0.97 | (0.88-1.06) | 0.97 | (0.88-1.06) | 1.07 | (0.97-1.17) | 1.06 | (0.97-1.17) |
| Normal HbA1c | 1.00 | ref | 1.00 | ref | 1.00 | ref | 1.00 | ref | 1.00 | ref |
| Pre-diabetes | 1.14 | (0.91-1.43) | 1.05 | (0.84-1.31) | 1.04 | (0.83-1.30) | 0.87 | (0.69-1.09) | 0.87 | (0.69-1.09) |
| Undiagnosed diabetes | 1.17 | (0.77-1.77) | 1.16 | (0.76-1.75) | 1.14 | (0.75-1.72) | 0.86 | (0.57-1.31) | 0.87 | (0.57-1.31) |
| Diagnosed diabetes | 1.44 | (1.23-1.69) | 1.28 | (1.09-1.50) | 1.27 | (1.08-1.49) | 1.02 | (0.87-1.20) | 1.06 | (0.89-1.25) |
| Women | | | | | | | | | | |
| Low-normal HbA1c | 0.66 | (0.59-0.73) | 0.82 | (0.74-0.91) | 0.82 | (0.74-0.92) | 0.94 | (0.84-1.04) | 0.93 | (0.84-1.04) |
| Normal HbA1c | 1.00 | ref | 1.00 | ref | 1.00 | ref | 1.00 | ref | 1.00 | ref |
| Pre-diabetes | 1.38 | (1.11-1.73) | 1.26 | (1.00-1.58) | 1.25 | (0.99-1.56) | 0.97 | (0.78-1.22) | 0.97 | (0.77-1.21) |
| Undiagnosed diabetes | 1.34 | (0.77-2.32) | 1.30 | (0.75-2.25) | 1.29 | (0.75-2.24) | 0.87 | (0.50-1.51) | 0.87 | (0.50-1.51) |
| Diagnosed diabetes | 1.56 | (1.25-1.94) | 1.44 | (1.16-1.80) | 1.42 | (1.14-1.77) | 0.96 | (0.77-1.21) | 0.99 | (0.78-1.24) |
| *p-interaction* | *0.0013* | | *0.0461* | | *0.0506* | | *0.3370* | | *0.3337* | |
| **Pulmonary embolism** | | | | | | | | | | |
| Men | | | | | | | | | | |
| Low-normal HbA1c | 0.76 | (0.70-0.82) | 0.88 | (0.81-0.95) | 0.88 | (0.81-0.95) | 0.98 | (0.91-1.06) | 0.99 | (0.92-1.07) |
| Normal HbA1c | 1.00 | ref | 1.00 | ref | 1.00 | ref | 1.00 | ref | 1.00 | ref |
| Pre-diabetes | 1.48 | (1.26-1.73) | 1.34 | (1.14-1.57) | 1.33 | (1.13-1.56) | 1.08 | (0.92-1.27) | 1.07 | (0.91-1.26) |
| Undiagnosed diabetes | 1.32 | (0.97-1.80) | 1.31 | (0.96-1.78) | 1.29 | (0.94-1.76) | 0.93 | (0.68-1.28) | 0.92 | (0.68-1.26) |
| Diagnosed diabetes | 1.37 | (1.20-1.56) | 1.21 | (1.06-1.37) | 1.19 | (1.05-1.36) | 0.92 | (0.81-1.05) | 0.99 | (0.86-1.14) |
| Women | | | | | | | | | | |
| Low-normal HbA1c | 0.60 | (0.56-0.65) | 0.77 | (0.71-0.83) | 0.77 | (0.71-0.83) | 0.88 | (0.81-0.96) | 0.89 | (0.82-0.97) |
| Normal HbA1c | 1.00 | ref | 1.00 | ref | 1.00 | ref | 1.00 | ref | 1.00 | ref |

| | | | | | | | | | | |
|---|---|---|---|---|---|---|---|---|---|---|
| Pre-diabetes | 1.53 | (1.30-1.80) | 1.38 | (1.17-1.63) | 1.37 | (1.16-1.61) | 1.05 | (0.89-1.24) | 1.03 | (0.87-1.22) |
| Undiagnosed diabetes | 1.12 | (0.71-1.76) | 1.08 | (0.69-1.70) | 1.07 | (0.68-1.69) | 0.70 | (0.44-1.10) | 0.69 | (0.44-1.08) |
| Diagnosed diabetes | 1.63 | (1.38-1.92) | 1.51 | (1.28-1.78) | 1.48 | (1.25-1.74) | 0.97 | (0.82-1.15) | 1.02 | (0.86-1.22) |
| *p-interaction* | *0.0001* | | *0.0110* | | *0.0133* | | *0.2768* | | *0.3009* | |
| **Stroke** | | | | | | | | | | |
| Men | | | | | | | | | | |
| Low-normal HbA1c | 0.71 | (0.67-0.76) | 0.87 | (0.82-0.93) | 0.88 | (0.83-0.93) | 0.95 | (0.89-1.01) | 0.98 | (0.92-1.04) |
| Normal HbA1c | 1.00 | ref | 1.00 | ref | 1.00 | ref | 1.00 | ref | 1.00 | ref |
| Pre-diabetes | 1.45 | (1.28-1.65) | 1.27 | (1.12-1.44) | 1.24 | (1.09-1.41) | 1.10 | (0.97-1.26) | 1.05 | (0.92-1.20) |
| Undiagnosed diabetes | 1.49 | (1.18-1.89) | 1.47 | (1.16-1.86) | 1.42 | (1.12-1.80) | 1.21 | (0.95-1.53) | 1.17 | (0.93-1.48) |
| Diagnosed diabetes | 2.14 | (1.96-2.33) | 1.80 | (1.65-1.96) | 1.75 | (1.60-1.91) | 1.58 | (1.44-1.73) | 1.42 | (1.29-1.56) |
| Women | | | | | | | | | | |
| Low-normal HbA1c | 0.66 | (0.61-0.70) | 0.91 | (0.85-0.98) | 0.92 | (0.86-0.99) | 0.99 | (0.92-1.06) | 1.01 | (0.94-1.08) |
| Normal HbA1c | 1.00 | ref | 1.00 | ref | 1.00 | ref | 1.00 | ref | 1.00 | ref |
| Pre-diabetes | 1.66 | (1.43-1.91) | 1.43 | (1.24-1.64) | 1.39 | (1.20-1.60) | 1.22 | (1.06-1.41) | 1.14 | (0.99-1.32) |
| Undiagnosed diabetes | 1.30 | (0.89-1.91) | 1.24 | (0.85-1.82) | 1.20 | (0.82-1.75) | 0.99 | (0.68-1.45) | 0.93 | (0.64-1.37) |
| Diagnosed diabetes | 2.28 | (2.00-2.59) | 2.02 | (1.77-2.30) | 1.93 | (1.70-2.20) | 1.62 | (1.42-1.85) | 1.43 | (1.25-1.64) |
| *p-interaction* | *0.1244* | | *0.3902* | | *0.4571* | | *0.6503* | | *0.6971* | |
| **Heart failure** | | | | | | | | | | |
| Men | | | | | | | | | | |
| Low-normal HbA1c | 0.62 | (0.59-0.65) | 0.78 | (0.74-0.82) | 0.79 | (0.75-0.83) | 0.92 | (0.87-0.98) | 0.99 | (0.94-1.05) |
| Normal HbA1c | 1.00 | ref | 1.00 | ref | 1.00 | ref | 1.00 | ref | 1.00 | ref |
| Pre-diabetes | 1.90 | (1.72-2.10) | 1.64 | (1.49-1.81) | 1.59 | (1.44-1.75) | 1.20 | (1.09-1.32) | 1.08 | (0.98-1.19) |
| Undiagnosed diabetes | 1.98 | (1.66-2.36) | 1.96 | (1.64-2.33) | 1.86 | (1.56-2.21) | 1.21 | (1.01-1.44) | 1.15 | (0.96-1.37) |
| Diagnosed diabetes | 3.01 | (2.82-3.21) | 2.50 | (2.34-2.67) | 2.39 | (2.23-2.55) | 1.69 | (1.58-1.81) | 1.27 | (1.18-1.36) |
| Women | | | | | | | | | | |
| Low-normal HbA1c | 0.60 | (0.56-0.65) | 0.88 | (0.82-0.94) | 0.89 | (0.82-0.95) | 1.04 | (0.97-1.12) | 1.08 | (1.01-1.17) |
| Normal HbA1c | 1.00 | ref | 1.00 | ref | 1.00 | ref | 1.00 | ref | 1.00 | ref |
| Pre-diabetes | 2.43 | (2.16-2.73) | 2.06 | (1.83-2.32) | 2.00 | (1.78-2.25) | 1.43 | (1.27-1.61) | 1.25 | (1.11-1.41) |
| Undiagnosed diabetes | 2.29 | (1.72-3.04) | 2.17 | (1.64-2.89) | 2.07 | (1.56-2.75) | 1.20 | (0.90-1.60) | 1.07 | (0.80-1.42) |
| Diagnosed diabetes | 4.15 | (3.76-4.58) | 3.66 | (3.31-4.03) | 3.43 | (3.11-3.79) | 1.98 | (1.79-2.19) | 1.44 | (1.29-1.60) |
| *p-interaction* | *<0.0001* | | *<0.0001* | | *<0.0001* | | *0.0124* | | *0.0740* | |

*A composite measure of all examined outcomes

*Abbreviations:* HbA1c, glycosylated haemoglobin; HR, hazard ratio; CI, confidence interval

*Notes:* Categories were defined by baseline HbA1c levels as follows: low-normal (<35 mmol/mol or <5.5%), normal (35-41 mmol/mol or 5.5-5.9%), pre-diabetes (42-47 mmol/mol or 6.0-6.4%), undiagnosed diabetes (≥48 mmol/mol or ≥6.5%), or diagnosed diabetes defined by a diagnostic code or receipt of glucose-lowering medication. Sex-specific hazard ratios from Cox proportional hazards models, adjusted for age at study entry, socio-demographics (i.e., ethnicity, deprivation), lifestyle characteristics (i.e., smoking status, alcohol consumption, physical activity, body mass index, waist-hip ratio, processed meat and fruit and vegetable intake), and clinical characteristics (i.e., total cholesterol, estimated glomerular filtration rate, C-reactive protein, diagnosed hypertension, use of antihypertensive medication or statins, and family history of cardiovascular disease).

**eTable 6.** Sensitivity analysis excluding individuals with diagnosed diabetes

| | Unadjusted | | Age-adjusted | | + Socio-demographics | | + Lifestyle characteristics | | + Clinical characteristics | |
|---|---|---|---|---|---|---|---|---|---|---|
| | **HR** | **(95% CI)** | **HR** | **(95% CI)** | **HR** | **(95% CI)** | **HR** | **(95% CI)** | **HR** | **(95% CI)** |
| **Any cardiovascular disease*** | | | | | | | | | | |
| Men | | | | | | | | | | |
| Low-normal HbA1c | 0.72 | (0.70-0.74) | 0.86 | (0.84-0.89) | 0.87 | (0.85-0.89) | 0.95 | (0.93-0.97) | 0.98 | (0.96-1.01) |
| Normal HbA1c | 1.00 | ref | 1.00 | ref | 1.00 | ref | 1.00 | ref | 1.00 | ref |
| Pre-diabetes | 1.46 | (1.38-1.54) | 1.3 | (1.23-1.37) | 1.29 | (1.22-1.36) | 1.09 | (1.03-1.15) | 1.04 | (0.98-1.10) |
| Undiagnosed diabetes | 1.32 | (1.19-1.46) | 1.31 | (1.18-1.45) | 1.29 | (1.16-1.43) | 1 | (0.90-1.11) | 0.98 | (0.88-1.09) |
| Women | | | | | | | | | | |
| Low-normal HbA1c | 0.64 | (0.62-0.65) | 0.87 | (0.84-0.89) | 0.87 | (0.84-0.90) | 0.96 | (0.93-0.99) | 0.99 | (0.96-1.02) |
| Normal HbA1c | 1.00 | ref | 1.00 | ref | 1.00 | ref | 1.00 | ref | 1.00 | ref |
| Pre-diabetes | 1.67 | (1.57-1.77) | 1.47 | (1.38-1.56) | 1.46 | (1.37-1.55) | 1.18 | (1.11-1.26) | 1.11 | (1.04-1.18) |
| Undiagnosed diabetes | 1.38 | (1.18-1.62) | 1.33 | (1.14-1.56) | 1.32 | (1.13-1.54) | 0.93 | (0.80-1.09) | 0.89 | (0.76-1.04) |
| *p-interaction* | *<0.0001* | | *0.0380* | | *0.0284* | | *0.1844* | | *0.2966* | |
| **Coronary artery disease** | | | | | | | | | | |
| Men | | | | | | | | | | |
| Low-normal HbA1c | 0.64 | (0.61-0.68) | 0.75 | (0.71-0.78) | 0.75 | (0.71-0.79) | 0.84 | (0.80-0.89) | 0.90 | (0.85-0.95) |
| Normal HbA1c | 1.00 | ref | 1.00 | ref | 1.00 | ref | 1.00 | ref | 1.00 | ref |
| Pre-diabetes | 1.58 | (1.43-1.75) | 1.44 | (1.30-1.59) | 1.42 | (1.28-1.57) | 1.22 | (1.10-1.35) | 1.16 | (1.05-1.29) |
| Undiagnosed diabetes | 1.76 | (1.48-2.10) | 1.74 | (1.46-2.07) | 1.70 | (1.43-2.03) | 1.41 | (1.18-1.68) | 1.35 | (1.13-1.61) |
| Women | | | | | | | | | | |
| Low-normal HbA1c | 0.56 | (0.52-0.60) | 0.71 | (0.65-0.77) | 0.71 | (0.66-0.77) | 0.82 | (0.76-0.89) | 0.89 | (0.82-0.96) |
| Normal HbA1c | 1.00 | ref | 1.00 | ref | 1.00 | ref | 1.00 | ref | 1.00 | ref |
| Pre-diabetes | 1.70 | (1.46-1.96) | 1.53 | (1.32-1.77) | 1.52 | (1.32-1.77) | 1.20 | (1.04-1.39) | 1.13 | (0.98-1.31) |
| Undiagnosed diabetes | 1.57 | (1.09-2.25) | 1.52 | (1.06-2.18) | 1.49 | (1.04-2.14) | 1.07 | (0.74-1.53) | 1.00 | (0.70-1.44) |
| *p-interaction* | *0.0092* | | *0.4835* | | *0.4419* | | *0.5523* | | *0.5083* | |
| **Atrial fibrillation** | | | | | | | | | | |
| Men | | | | | | | | | | |
| Low-normal HbA1c | 0.75 | (0.72-0.77) | 0.95 | (0.91-0.98) | 0.94 | (0.91-0.98) | 1.02 | (0.98-1.06) | 1.04 | (1.00-1.08) |
| Normal HbA1c | 1.00 | ref | 1.00 | ref | 1.00 | ref | 1.00 | ref | 1.00 | ref |
| Pre-diabetes | 1.50 | (1.40-1.62) | 1.29 | (1.20-1.40) | 1.3 | (1.21-1.41) | 1.09 | (1.01-1.18) | 1.03 | (0.96-1.12) |
| Undiagnosed diabetes | 1.37 | (1.18-1.58) | 1.35 | (1.17-1.56) | 1.36 | (1.18-1.57) | 1.03 | (0.89-1.19) | 1.02 | (0.88-1.18) |
| Women | | | | | | | | | | |
| Low-normal HbA1c | 0.67 | (0.64-0.70) | 0.99 | (0.94-1.04) | 0.99 | (0.94-1.03) | 1.06 | (1.02-1.12) | 1.06 | (1.01-1.11) |
| Normal HbA1c | 1.00 | ref | 1.00 | ref | 1.00 | ref | 1.00 | ref | 1.00 | ref |
| Pre-diabetes | 1.59 | (1.45-1.74) | 1.35 | (1.22-1.48) | 1.36 | (1.24-1.50) | 1.12 | (1.02-1.23) | 1.04 | (0.95-1.15) |
| Undiagnosed diabetes | 1.29 | (1.01-1.65) | 1.22 | (0.95-1.56) | 1.25 | (0.98-1.60) | 0.9 | (0.70-1.15) | 0.85 | (0.66-1.09) |
| *p-interaction* | *0.0006* | | *0.3974* | | *0.4168* | | *0.3750* | | *0.5336* | |
| **Deep vein thrombosis** | | | | | | | | | | |
| Men | | | | | | | | | | |
| Low-normal HbA1c | 0.85 | (0.77-0.93) | 0.97 | (0.88-1.06) | 0.97 | (0.89-1.07) | 1.07 | (0.98-1.18) | 1.07 | (0.97-1.18) |
| Normal HbA1c | 1.00 | ref | 1.00 | ref | 1.00 | ref | 1.00 | ref | 1.00 | ref |
| Pre-diabetes | 1.22 | (0.98-1.51) | 1.11 | (0.89-1.38) | 1.1 | (0.89-1.37) | 0.91 | (0.73-1.14) | 0.91 | (0.73-1.14) |
| Undiagnosed diabetes | 1.14 | (0.76-1.73) | 1.13 | (0.75-1.71) | 1.12 | (0.74-1.69) | 0.84 | (0.55-1.27) | 0.84 | (0.55-1.27) |
| Women | | | | | | | | | | |
| Low-normal HbA1c | 0.67 | (0.60-0.74) | 0.84 | (0.75-0.93) | 0.84 | (0.76-0.93) | 0.95 | (0.86-1.06) | 0.95 | (0.85-1.05) |
| Normal HbA1c | 1.00 | ref | 1.00 | ref | 1.00 | ref | 1.00 | ref | 1.00 | ref |
| Pre-diabetes | 1.47 | (1.18-1.83) | 1.34 | (1.08-1.66) | 1.32 | (1.06-1.64) | 1.03 | (0.83-1.28) | 1.02 | (0.82-1.27) |
| Undiagnosed diabetes | 1.52 | (0.91-2.53) | 1.47 | (0.88-2.45) | 1.46 | (0.88-2.43) | 0.98 | (0.59-1.64) | 0.98 | (0.58-1.63) |
| *p-interaction* | *0.0011* | | *0.0473* | | *0.0491* | | *0.2250* | | *0.2303* | |
| **Pulmonary embolism** | | | | | | | | | | |
| Men | | | | | | | | | | |
| Low-normal HbA1c | 0.76 | (0.70-0.82) | 0.88 | (0.82-0.95) | 0.88 | (0.82-0.95) | 0.99 | (0.91-1.06) | 1 | (0.92-1.08) |
| Normal HbA1c | 1.00 | ref | 1.00 | ref | 1.00 | ref | 1.00 | ref | 1.00 | ref |
| Pre-diabetes | 1.47 | (1.25-1.72) | 1.33 | (1.14-1.56) | 1.32 | (1.13-1.55) | 1.07 | (0.91-1.26) | 1.07 | (0.91-1.25) |
| Undiagnosed diabetes | 1.33 | (0.98-1.81) | 1.32 | (0.97-1.79) | 1.30 | (0.96-1.77) | 0.94 | (0.69-1.28) | 0.93 | (0.68-1.26) |
| Women | | | | | | | | | | |
| Low-normal HbA1c | 0.61 | (0.56-0.66) | 0.78 | (0.72-0.85) | 0.78 | (0.72-0.85) | 0.9 | (0.83-0.98) | 0.91 | (0.84-0.99) |
| Normal HbA1c | 1.00 | ref | 1.00 | ref | 1.00 | ref | 1.00 | ref | 1.00 | ref |
| Pre-diabetes | 1.59 | (1.36-1.87) | 1.44 | (1.23-1.69) | 1.42 | (1.21-1.67) | 1.09 | (0.92-1.28) | 1.07 | (0.91-1.26) |
| Undiagnosed diabetes | 1.17 | (0.75-1.81) | 1.13 | (0.72-1.75) | 1.12 | (0.72-1.74) | 0.73 | (0.47-1.13) | 0.71 | (0.46-1.11) |
| *p-interaction* | *0.0007* | | *0.1135* | | *0.1139* | | *0.3127* | | *0.3135* | |
| **Stroke** | | | | | | | | | | |
| Men | | | | | | | | | | |

| | | | | | | | | | | |
|---|---|---|---|---|---|---|---|---|---|---|
| Low-normal HbA1c | 0.71 | (0.67-0.75) | 0.87 | (0.82-0.93) | 0.88 | (0.83-0.93) | 0.95 | (0.89-1.01) | 0.98 | (0.92-1.04) |
| Normal HbA1c | 1.00 | ref | 1.00 | ref | 1.00 | ref | 1.00 | ref | 1.00 | ref |
| Pre-diabetes | 1.44 | (1.27-1.64) | 1.26 | (1.10-1.43) | 1.23 | (1.08-1.40) | 1.1 | (0.96-1.25) | 1.04 | (0.92-1.19) |
| Undiagnosed diabetes | 1.52 | (1.21-1.91) | 1.50 | (1.19-1.88) | 1.45 | (1.15-1.83) | 1.24 | (0.98-1.56) | 1.20 | (0.95-1.52) |
| Women | | | | | | | | | | |
| Low-normal HbA1c | 0.65 | (0.61-0.70) | 0.92 | (0.85-0.98) | 0.92 | (0.86-0.99) | 0.99 | (0.92-1.06) | 1.01 | (0.94-1.08) |
| Normal HbA1c | 1.00 | ref | 1.00 | ref | 1.00 | ref | 1.00 | ref | 1.00 | ref |
| Pre-diabetes | 1.65 | (1.44-1.91) | 1.42 | (1.23-1.63) | 1.39 | (1.20-1.60) | 1.23 | (1.06-1.41) | 1.15 | (0.99-1.33) |
| Undiagnosed diabetes | 1.33 | (0.91-1.93) | 1.26 | (0.87-1.83) | 1.22 | (0.84-1.78) | 1.02 | (0.70-1.49) | 0.96 | (0.66-1.40) |
| *p-interaction* | | *0.1015* | | *0.3903* | | *0.4020* | | *0.4386* | | *0.4923* |
| **Heart failure** | | | | | | | | | | |
| Men | | | | | | | | | | |
| Low-normal HbA1c | 0.62 | (0.59-0.66) | 0.79 | (0.74-0.83) | 0.8 | (0.75-0.84) | 0.93 | (0.88-0.98) | 1 | (0.94-1.06) |
| Normal HbA1c | 1.00 | ref | 1.00 | ref | 1.00 | ref | 1.00 | ref | 1.00 | ref |
| Pre-diabetes | 1.91 | (1.73-2.11) | 1.65 | (1.49-1.81) | 1.59 | (1.44-1.76) | 1.2 | (1.09-1.32) | 1.08 | (0.97-1.19) |
| Undiagnosed diabetes | 1.99 | (1.67-2.36) | 1.96 | (1.65-2.33) | 1.86 | (1.56-2.22) | 1.2 | (1.00-1.43) | 1.15 | (0.96-1.37) |
| Women | | | | | | | | | | |
| Low-normal HbA1c | 0.60 | (0.56-0.65) | 0.89 | (0.83-0.95) | 0.9 | (0.83-0.96) | 1.05 | (0.98-1.13) | 1.09 | (1.02-1.18) |
| Normal HbA1c | 1.00 | ref | 1.00 | ref | 1.00 | ref | 1.00 | ref | 1.00 | ref |
| Pre-diabetes | 2.43 | (2.16-2.73) | 2.05 | (1.83-2.31) | 1.99 | (1.77-2.24) | 1.41 | (1.25-1.59) | 1.24 | (1.10-1.40) |
| Undiagnosed diabetes | 2.27 | (1.71-3.01) | 2.15 | (1.62-2.86) | 2.05 | (1.55-2.73) | 1.17 | (0.88-1.56) | 1.04 | (0.78-1.38) |
| *p-interaction* | | *0.0080* | | *0.0064* | | *0.0064* | | *0.0211* | | *0.0840* |

*A composite measure of all examined outcomes

*Abbreviations:* HbA1c, glycosylated haemoglobin; HR, hazard ratio; CI, confidence interval

*Notes:* Categories were defined by baseline HbA1c levels as follows: low-normal (<35 mmol/mol or <5.5%), normal (35-41 mmol/mol or 5.5-5.9%), pre-diabetes (42-47 mmol/mol or 6.0-6.4%), or undiagnosed diabetes (≥48 mmol/mol or ≥6.5%). Sex-specific hazard ratios from Cox proportional hazards models, adjusted for age at study entry, socio-demographics (i.e., ethnicity, deprivation), lifestyle characteristics (i.e., smoking status, alcohol consumption, physical activity, body mass index, waist-hip ratio, processed meat and fruit and vegetable intake), and clinical characteristics (i.e., total cholesterol, estimated glomerular filtration rate, C-reactive protein, diagnosed hypertension, use of antihypertensive medication or statins, and family history of cardiovascular disease).

**eFigure 1.** Schoenfeld residuals testing the proportional hazards assumption between HbA1c category and any cardiovascular disease, by sex

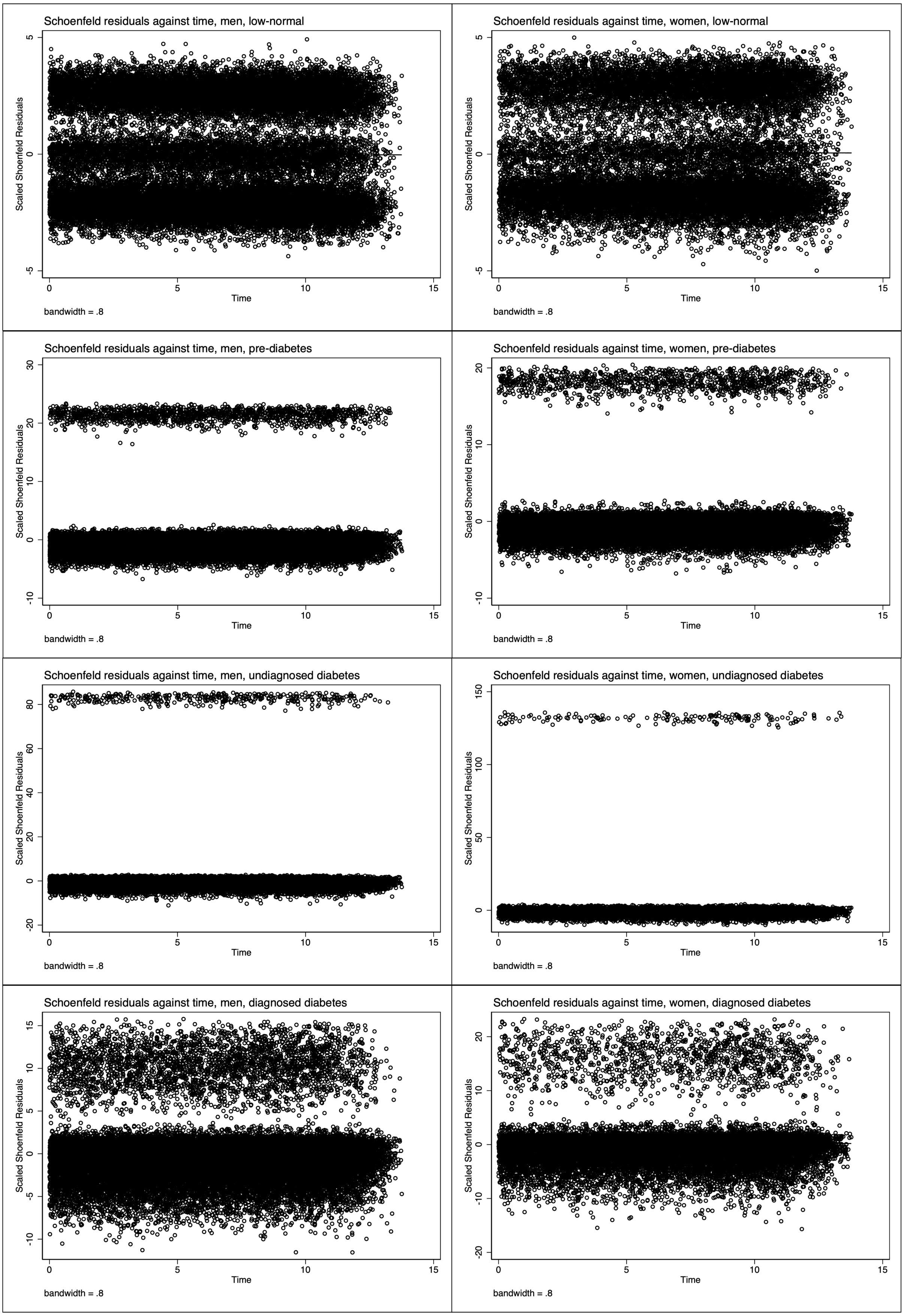

**The RECORD statement – checklist of items, extended from the STROBE statement, that should be reported in observational studies using routinely collected health data**

| | Item No. | STROBE items | Location in manuscript where items are reported | RECORD items | Location in manuscript where items are reported |
|---|---|---|---|---|---|
| **Title and abstract** | | | | | |
| | 1 | (a) Indicate the study's design with a commonly used term in the title or the abstract (b) Provide in the abstract an informative and balanced summary of what was done and what was found | (a) Title & Abstract<br><br>(b) Abstract | RECORD 1.1: The type of data used should be specified in the title or abstract. When possible, the name of the databases used should be included.<br><br>RECORD 1.2: If applicable, the geographic region and timeframe within which the study took place should be reported in the title or abstract.<br><br>RECORD 1.3: If linkage between databases was conducted for the study, this should be clearly stated in the title or abstract. | 1.1: Abstract – Methods<br><br>1.2: Abstract – Methods & Findings<br><br>1.3: Abstract – Methods |
| **Introduction** | | | | | |
| Background rationale | 2 | Explain the scientific background and rationale for the investigation being reported | Background (Para 1 & 2) | | |
| Objectives | 3 | State specific objectives, including any prespecified hypotheses | Background (End of Para 3) | | |
| **Methods** | | | | | |
| Study Design | 4 | Present key elements of study design early in the paper | Methods (Study design and population) | | |
| Setting | 5 | Describe the setting, locations, and relevant dates, including periods of recruitment, exposure, follow-up, and data collection | Methods (Study design and population; Exposure, | | |

| | | | | | |
|---|---|---|---|---|---|
| | | | outcome and follow-up) | | |
| Participants | 6 | *(a) Cohort study* - Give the eligibility criteria, and the sources and methods of selection of participants. Describe methods of follow-up<br>*Case-control study* - Give the eligibility criteria, and the sources and methods of case ascertainment and control selection. Give the rationale for the choice of cases and controls<br>*Cross-sectional study* - Give the eligibility criteria, and the sources and methods of selection of participants<br><br>*(b) Cohort study* - For matched studies, give matching criteria and number of exposed and unexposed<br>*Case-control study* - For matched studies, give matching criteria and the number of controls per case | (a) Methods (Study design and population; Exposure, outcome and follow-up) | RECORD 6.1: The methods of study population selection (such as codes or algorithms used to identify subjects) should be listed in detail. If this is not possible, an explanation should be provided.<br><br>RECORD 6.2: Any validation studies of the codes or algorithms used to select the population should be referenced. If validation was conducted for this study and not published elsewhere, detailed methods and results should be provided.<br><br>RECORD 6.3: If the study involved linkage of databases, consider use of a flow diagram or other graphical display to demonstrate the data linkage process, including the number of individuals with linked data at each stage. | 6.1: Methods (Study design and population; Exposure, outcome and follow-up)<br><br>6.2: Methods (Exposure, outcome and follow-up; Covariates) & Supplementary Appendix<br><br>6.3: Figure 1 |
| Variables | 7 | Clearly define all outcomes, exposures, predictors, potential confounders, and effect modifiers. Give diagnostic criteria, if applicable. | Methods (Exposure, outcome and follow-up; Covariates) | RECORD 7.1: A complete list of codes and algorithms used to classify exposures, outcomes, confounders, and effect modifiers should be provided. If these cannot be reported, an explanation should be provided. | 7.1: Methods (Exposure, outcome and follow-up; Covariates) & Supplementary Appendix |
| Data sources/ measurement | 8 | For each variable of interest, give sources of data and details of methods of assessment (measurement). | Methods (Exposure, outcome and follow-up; Covariates). | | |

| | | | | | |
|---|---|---|---|---|---|
| | | Describe comparability of assessment methods if there is more than one group | | | |
| Bias | 9 | Describe any efforts to address potential sources of bias | Methods (Covariates; Statistical analysis; Sensitivity analyses) | | |
| Study size | 10 | Explain how the study size was arrived at | Methods (all) | | |
| Quantitative variables | 11 | Explain how quantitative variables were handled in the analyses. If applicable, describe which groupings were chosen, and why | Methods (Exposure, outcome and follow-up; Statistical analysis) | | |
| Statistical methods | 12 | (a) Describe all statistical methods, including those used to control for confounding<br>(b) Describe any methods used to examine subgroups and interactions<br>(c) Explain how missing data were addressed<br>(d) *Cohort study* - If applicable, explain how loss to follow-up was addressed<br>*Case-control study* - If applicable, explain how matching of cases and controls was addressed<br>*Cross-sectional study* - If applicable, describe analytical methods taking account of sampling strategy<br>(e) Describe any sensitivity analyses | (a-c) Methods (Covariates; Statistical analysis; Sensitivity analyses)<br><br>(d) Not applicable for this study<br><br>(e) Methods (Sensitivity analyses) | | |
| Data access and cleaning methods | | .. | | RECORD 12.1: Authors should describe the extent to which the investigators had access to the database population used to create the study population. | 12.1: Methods (Role of the funding source)<br><br>12.2: Methods (Study design and |

| | | | | | |
|---|---|---|---|---|---|
| | | | | RECORD 12.2: Authors should provide information on the data cleaning methods used in the study. | population; Covariates) |
| Linkage | | .. | | RECORD 12.3: State whether the study included person-level, institutional-level, or other data linkage across two or more databases. The methods of linkage and methods of linkage quality evaluation should be provided. | Methods (Study design and population) |
| **Results** | | | | | |
| Participants | 13 | (a) Report the numbers of individuals at each stage of the study (*e.g.*, numbers potentially eligible, examined for eligibility, confirmed eligible, included in the study, completing follow-up, and analysed)<br>(b) Give reasons for non-participation at each stage.<br>(c) Consider use of a flow diagram | (a-c) Results (Cohort description; Figure 1) | RECORD 13.1: Describe in detail the selection of the persons included in the study (*i.e.,* study population selection) including filtering based on data quality, data availability and linkage. The selection of included persons can be described in the text and/or by means of the study flow diagram. | 13.1: Results (Cohort description; Figure 1) |
| Descriptive data | 14 | (a) Give characteristics of study participants (*e.g.*, demographic, clinical, social) and information on exposures and potential confounders<br>(b) Indicate the number of participants with missing data for each variable of interest<br>(c) *Cohort study* - summarise follow-up time (*e.g.*, average and total amount) | (a) Results (Cohort description; Table 1)<br><br>(b) Methods (Covariates); Table 1<br><br>(c) Results (Age-standardised incidence rates) | | |
| Outcome data | 15 | *Cohort study* - Report numbers of outcome events or summary measures over time<br>*Case-control study* - Report numbers in each exposure category, or summary measures of exposure | Results (Age-standardised incidence rates) | | |

| | | | | | |
|---|---|---|---|---|---|
| | | *Cross-sectional study* - Report numbers of outcome events or summary measures | | | |
| Main results | 16 | (a) Give unadjusted estimates and, if applicable, confounder-adjusted estimates and their precision (e.g., 95% confidence interval). Make clear which confounders were adjusted for and why they were included<br>(b) Report category boundaries when continuous variables were categorized<br>(c) If relevant, consider translating estimates of relative risk into absolute risk for a meaningful time period | (a) Results (Figure 2; eTable 2)<br><br>(b) Results (all)<br><br>(c) Results (Age-standardised incidence rates) | | |
| Other analyses | 17 | Report other analyses done—e.g., analyses of subgroups and interactions, and sensitivity analyses | Results (HbA1c category and CVD risk; Identifying factors most responsible for attenuating excess risk; Sensitivity analyses) | | |
| **Discussion** | | | | | |
| Key results | 18 | Summarise key results with reference to study objectives | Discussion (para 1) | | |
| Limitations | 19 | Discuss limitations of the study, taking into account sources of potential bias or imprecision. Discuss both direction and magnitude of any potential bias | Discussion (para 6) | RECORD 19.1: Discuss the implications of using data that were not created or collected to answer the specific research question(s). Include discussion of misclassification bias, unmeasured confounding, missing data, and changing eligibility over time, as they pertain to the study being reported. | 19.1 Discussion (para 6) |
| Interpretation | 20 | Give a cautious overall interpretation of results considering objectives, limitations, multiplicity of | Discussion (Throughout) | | |

| | | | | | |
|---|---|---|---|---|---|
| | | analyses, results from similar studies, and other relevant evidence | | | |
| Generalisability | 21 | Discuss the generalisability (external validity) of the study results | Discussion (para 6) | | |
| **Other Information** | | | | | |
| Funding | 22 | Give the source of funding and the role of the funders for the present study and, if applicable, for the original study on which the present article is based | Methods (Role of the funding source); Funding statement | | |
| Accessibility of protocol, raw data, and programming code | | .. | | RECORD 22.1: Authors should provide information on how to access any supplemental information such as the study protocol, raw data, or programming code. | Data availability statement |
